# Antidepressant Maintenance Versus Active Monitoring After Depression Remission: A Decision Analysis Stratified by Relapse Risk and Patient Preferences

**DOI:** 10.64898/2026.07.17.26358340

**Authors:** William U. Meyerson, Tianxi Cai, Jordan W. Smoller

**Affiliations:** Center for Precision Psychiatry, Massachusetts General Hospital, Boston, Massachusetts; Department of Psychiatry, Massachusetts General Hospital/Harvard Medical School, Boston, Massachusetts; Department of Biomedical Informatics, Harvard Medical School, Boston, Massachusetts; Department of Biostatistics, Harvard T.H. Chan School of Public Health, Boston, Massachusetts; Psychiatric and Neurodevelopmental Genetics Unit, Center for Genomic Medicine, Massachusetts General Hospital, Boston, Massachusetts

## Abstract

**Importance:** Patients who achieve remission from major depressive disorder (MDD) often face a preference-sensitive decision between continued antidepressant maintenance and discontinuation with active monitoring. Quantifying the tradeoff between depression burden and long-term medication exposure may support more individualized shared decision-making.

**Objective:** To quantify tradeoffs between continuous antidepressant maintenance and active monitoring after MDD remission, and to identify preference thresholds favoring each strategy across relapse-risk strata.

**Design:** Individual-level decision-analytic health-state transition model calibrated to randomized maintenance-discontinuation trials and a longitudinal first depressive episode cohort, with a 5-year time horizon.

**Setting:** Outpatient clinical decision after completion of an 8-month continuation phase following remission from MDD.

**Participants:** Adults in remission from MDD, represented across 4 clinically anchored relapse-risk strata ranging from very low risk after a first mild episode to high risk after highly recurrent depression.

**Exposures:** Continuous antidepressant maintenance vs discontinuation with active monitoring and antidepressant restart after detected relapse.

**Main Outcomes and Measures:** Severity-weighted depression-months, antidepressant medication-years, medication-years per depression-month averted, and net benefit across preference thresholds defined as the maximum additional medication-years a patient would be willing to accept to avert 1 depression-month.

**Results:** Continuous maintenance reduced depression burden but required substantially more medication exposure, with efficiency strongly dependent on relapse risk. Medication-years per depression-month averted ranged from 11.8 (95% uncertainty interval [UI], 7.8-19.6) in the very low-risk group to 1.5 (95% UI, 0.8-3.0) in the high-risk group. At a preference threshold of 3 medication-years per depression-month averted, maintenance was preferred for moderate- and high-risk patients; at a threshold of 2, only for high-risk patients; and at a threshold of 1, for no risk group.

**Conclusions and Relevance:** In this decision-analytic model, the value of continuous antidepressant maintenance depended strongly on baseline relapse risk and patient preferences regarding long-term medication exposure. These findings provide a quantitative framework for shared decision-making about antidepressant maintenance after remission from MDD.

## Introduction

Major depressive disorder (MDD) affects 1 in 5 adults over the life course ^1^, and relapse after remission is common and can be disabling ^2^. Patients in remission on antidepressants face a key decision: whether to continue maintenance treatment or discontinue medication with symptom monitoring and planned restart if symptoms return (active monitoring). Maintenance lowers relapse risk ^3,4^, but may bring burdens such as sexual side effects, weight gain, emotional numbing, and an unwanted sense of dependence on medication ^5– 7,35^. Although relapse-prevention psychotherapy is an effective option for some patients ^8^, access and acceptability vary ^9,10^, and routine decisions often still center on maintenance versus active monitoring ^11^.

Clinical guidelines on antidepressant maintenance are relatively straightforward for patients at the extremes of relapse risk ^12–15^. Long-term maintenance is generally discouraged for patients at lower risk and more strongly recommended for those at higher risk. Between these extremes, however, many patients have moderate relapse risk ^16^ and meaningful treatment burden ^5^. For these patients, the practical question is how much additional antidepressant exposure is worth accepting to reduce future time spent depressed.

Informed shared decisions require that patients understand relevant tradeoffs and relate them to their own values ^17^. Randomized maintenance-discontinuation trials and meta-analyses have reported that continued maintenance reduces 1-year relapse risk from roughly 40% to 20% ^3,4,18^. However, relapse is a coarse endpoint; decision-making also depends on time spent well versus unwell over months to years ^19^. These trials are typically enriched for recurrent illness (a mean of 4–5 prior episodes)^3^, which may limit generalizability to patients with more moderate baseline risk. Prior decision-analytic models have focused largely on cost-effectiveness rather than clinical tradeoffs for shared decision-making ^20–22^. Existing evidence thus does not provide a practical framework for judging whether continued antidepressants are likely to be value-concordant, given a patient’s relapse-risk level and treatment burden.

To address this gap, we developed a decision-analytic health-state transition model (microsimulation) projecting outcomes under continuous maintenance versus active monitoring for patients with MDD in remission. The model integrates trial and longitudinal cohort evidence and builds on our prior modeling work^23^. We express the core tradeoff as medication-years per depression-month averted, stratified by relapse-risk group. This follows a standard decision-analytic principle: preferences over long-term courses can be represented by the relative value of time spent in different health states ^24^. Patient preference thresholds then determine when that tradeoff favors maintenance versus active monitoring. In the present analysis, the tradeoff varied nearly eightfold: maintenance required approximately 1.5 additional years of antidepressant exposure to avert 1 month of depression in the highest-risk group, compared with nearly 12 years in the lowest-risk group. These estimates are intended to help clinicians and patients weigh relapse prevention against the patient’s valuation of long-term antidepressant exposure.

## Methods

### Overview

We developed an individual-level health state transition model (microsimulation) to inform antidepressant continuation decisions after remission from a major depressive episode. The simulated decision point was completion of an 8-month continuation phase after remission, when a patient and clinician may consider either ongoing antidepressant maintenance or guided discontinuation with active monitoring. The model compared continuous maintenance with guided discontinuation and planned antidepressant restart after detected relapse over a 5-year horizon.

The model was designed to estimate a clinical tradeoff across relapse-risk strata and preference thresholds, not deterministic patient-level predictions. The model includes depressive health states and transitions among them, including remission, relapse into an active major depressive episode, and recovery after relapse (Supplemental Figure S1).

We followed CHEERS 2022 guidance where applicable ^25^. Analyses used R, version 4.4.1 (R Foundation for Statistical Computing). The Mass General Brigham Institutional Review Board determined that this work did not constitute human subjects research due to reliance on aggregate data. The analysis plan was not preregistered.

### Treatment Strategies and Decision Structure

We compared two treatment strategies, illustrated in Figure 1. Under continuous maintenance, patients remained on antidepressants throughout follow-up. Under active monitoring, patients discontinued antidepressants at model entry after completing continuation treatment for the index episode; upon detected relapse, they restarted antidepressants and continued treatment for 8 months after remission before discontinuing again, consistent with guideline-recommended continuation periods in patients not pursuing maintenance treatment ^12–15^. The implied taper schedule in active monitoring reflects empirically the balance of tapering strategies used in the underlying RCTs, which is not consistently reported.

**Figure 1.**
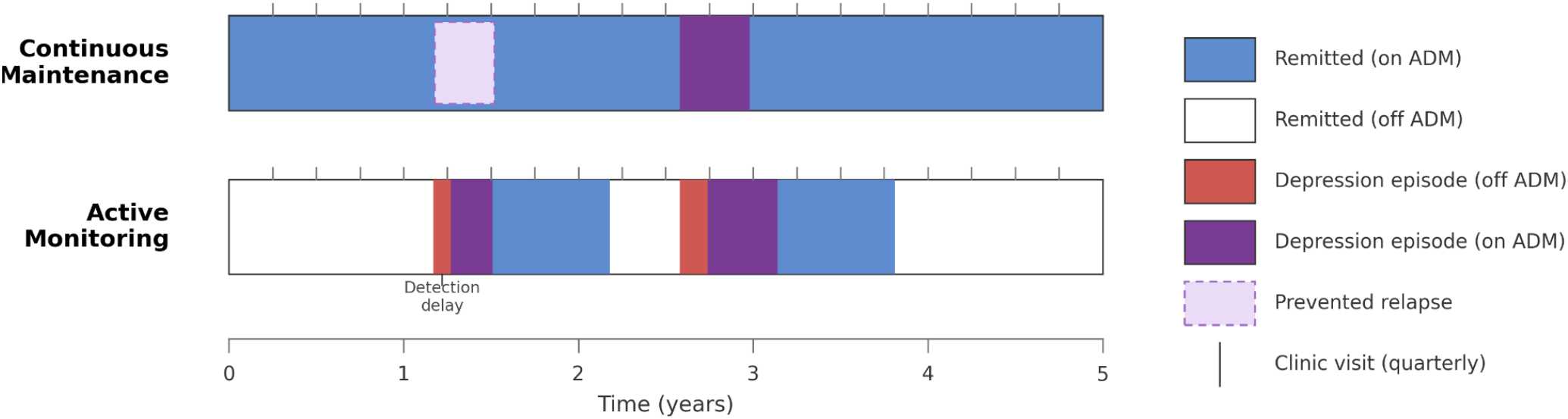
Schematic timelines for continuous maintenance and active monitoring over 5 years. Bars depict periods on antidepressants versus off treatment, with relapse-triggered restart and fixed continuation periods after remission; monitoring determines the delay between relapse onset and restart under active monitoring. A 1-month ramp-up period after treatment restart is not shown.

The model used a 1-month cycle length. Relapse-detection delay depended on clinical monitoring frequency, which was quarterly in the base case; sensitivity analyses examined monthly and every-6-month monitoring. The 5-year horizon was chosen to capture longer-term tradeoffs while remaining within the longest follow-up available in the source studies ^26^. We did not discount outcomes because both burdens were measured as patient-centered time accruing over the same bounded horizon.

### Overview of Model Inputs

For model inputs, we selected sources to supply complementary quantities that no single study reported across the full decision horizon. Kishi et al. (2023), a systematic review and meta-analysis of 35 double-blind maintenance-discontinuation randomized trials involving 9,442 adults with recurrent depression, was chosen because it provided the strongest randomized evidence on maintenance versus discontinuation and reported relapse estimates across the post-randomization trajectory, including cumulative 12-month relapse rates of approximately 44% after discontinuation and 22% with maintenance. These trials primarily represented highly recurrent populations, with a mean of approximately 4–5 prior depressive episodes. To extend the model toward patients with lower recurrence risk and longer follow-up, we used Bukh et al. (2016), a prospective study of 301 Danish patients receiving inpatient or outpatient specialty psychiatric care after a first lifetime depressive episode. It reported cumulative recurrence after remission of 9.0% at 1 year and 31.5% at 5 years. Selected model parameters and key structural assumptions are summarized in Table 1; full derivations are provided in the Supplement.

**Table 1.**
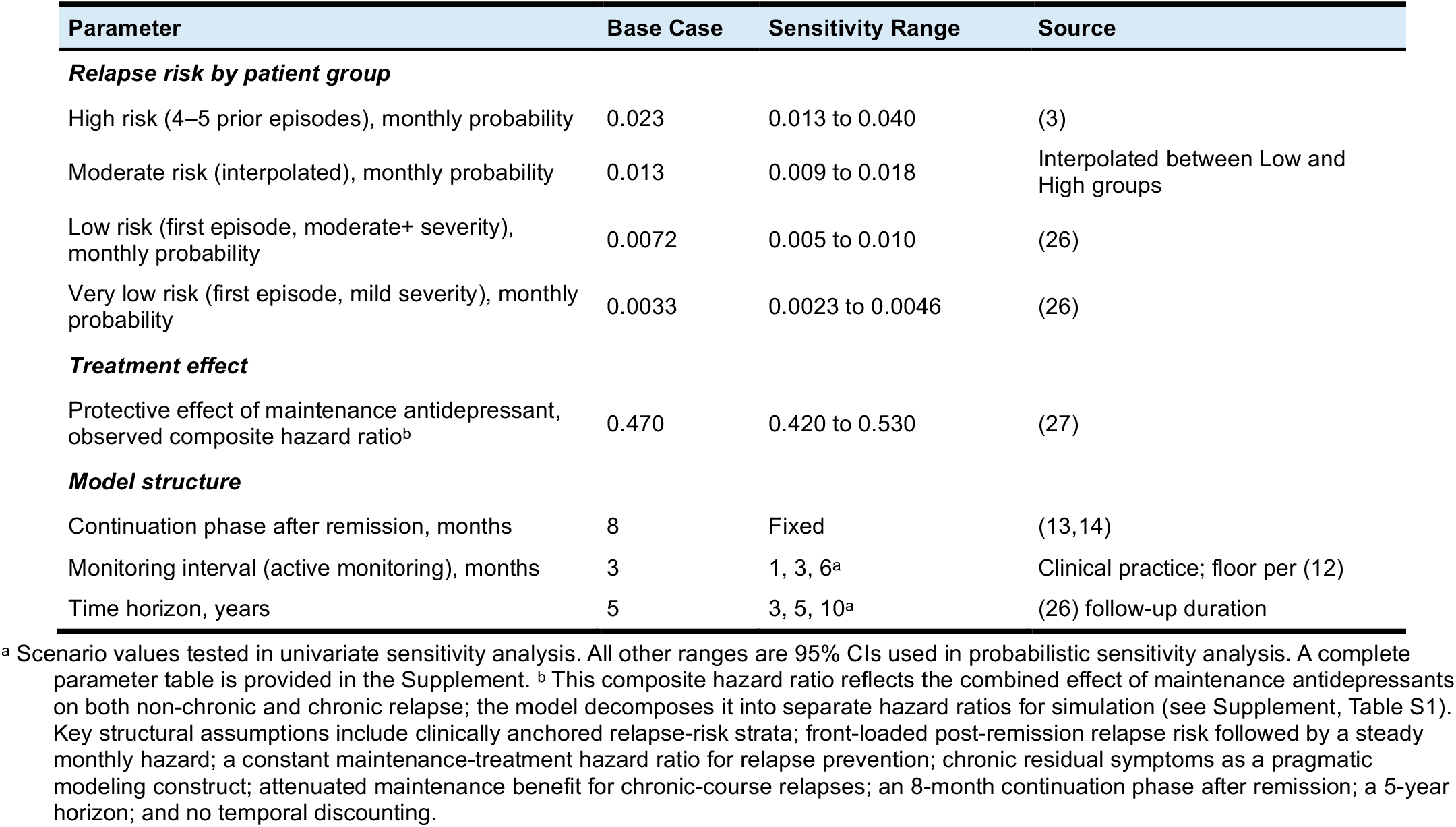
Selected model parameters.

### Outcomes and Preference Thresholds

The primary outcomes were depression-months and medication-years over 5 years. Depression-months were severity-weighted, with acute major depressive episodes counting fully and chronic residual symptoms contributing a lower weight derived from long-term cohort data ^19^. For each relapse-risk stratum, we report absolute outcomes under each strategy, between-strategy differences, and medication-years per depression-month averted, defined as patient-years of antidepressant exposure required to avert 1 patient-month of depression.

We also computed the net benefit of continuous maintenance versus active monitoring across preference thresholds for antidepressant exposure, defined as the maximum additional medication-years a patient would accept to avert 1 depression-month. These thresholds are hypothetical preference parameters, not empirically measured patient preferences. They represent how a patient values time spent depressed relative to side effects, functioning, quality of life, and other burdens of continued treatment.

The break-even threshold for each risk stratum equals its medication-years per depression-month averted. For example, if maintenance requires 2 additional medication-years per depression-month averted, patients willing to accept that exposure would prefer maintenance; others would prefer active monitoring. The primary outcome is net benefit across the full range of continuous thresholds. For illustration purposes only, we highlight thresholds of 1, 2, and 3 medication-years per depression-month averted as illustrative anchors to showcase different parts of the decision curve.

### Model Structure and Health States

At model entry, patients were in remission after completing continuation treatment. During monthly cycles, they could remain in remission, relapse, remit, or enter and later recover from chronic residual symptoms. Active major depressive episode represented full depressive morbidity after relapse; remission represented time without active major depressive episode or chronic residual symptoms. The chronic residual-symptom state captures persistent, lower-severity depressive morbidity after the acute phase of some episodes and is not equivalent to persistent depressive disorder or dysthymia.

### Relapse Risk and Maintenance Effect

Relapse risk was modeled to reflect baseline heterogeneity in recurrence propensity, higher risk soon after remission, and risk reduction during antidepressant maintenance. Complementary empirical anchors were needed because no single source spans both individualized relapse risk and the maintenance-versus-discontinuation contrast.

Each month, patients in remission faced a relapse probability that depended on baseline risk stratum, time since the most recent remission, and antidepressant status. The four relapse-risk strata were clinically anchored benchmarks rather than exhaustive patient categories or individualized predictions. The Very low and Low strata were anchored to a prospective first-lifetime depressive episode cohort stratified by baseline severity ^26^: Very low corresponded to the mild subgroup and Low to the subgroup with at least moderate baseline severity. The High-risk stratum was anchored to a maintenance-discontinuation trial meta-analysis in recurrent depression ^3^, representing populations with mean lifetime depressive episode counts of approximately 4 to 5. The Moderate stratum was interpolated between Low and High, notionally corresponding to about 2 to 3 lifetime depressive episodes. Monthly hazards were calibrated to cumulative relapse or recurrence in these sources.

Consistent with maintenance-discontinuation trial patterns, relapse hazards were elevated during the first 8 months after remission and lower thereafter. The time-varying relapse shape was derived from the same maintenance-discontinuation meta-analysis used for the High-risk anchor ^3^. Maintenance effects were modeled separately from baseline relapse hazard on the hazard scale, using a meta-analysis of relapse-prevention hazard ratios ^27^.

### Remission After Relapse

Once in an active depressive episode, patients faced monthly remission probabilities that depended on antidepressant status. Remission probabilities and antidepressant effects on remission were modeled on the hazard scale and calibrated to 6-week remission rates from an acute-phase trial meta-analysis ^28^. To reflect the observed time course of antidepressant benefit after treatment initiation or restart, treatment effects were modeled as partially active for 1 month and fully active thereafter, informed by a focused meta-analysis of antidepressant response dynamics ^29^.

### Chronic Residual Symptoms

To represent persistent depressive morbidity over longer horizons, the model allowed a subset of episodes to follow a chronic course. Chronic episode persistence was calibrated to long-term episode-duration data from a population-based cohort study ^30^, under a fixed on/off antidepressant exposure mixture reflecting treatment uptake in population-based cohorts ^31^. Relative daily burden during chronic residual symptoms compared with acute major depressive episode was estimated from a long-term cohort study ^19^.

In the base case, maintenance treatment was assumed to have half the relapse-prevention effect in chronic-course trajectories as in nonchronic episodes. Sensitivity analyses examined alternatives in which maintenance had either no relapse-prevention benefit or the same benefit as in nonchronic episodes.

### Model Verification and Validation

We evaluated model performance by simulating 10,000 patients per scenario and comparing model-projected outputs with published benchmarks across four domains: relapse after remission in maintenance-discontinuation trials ^3^, longer-horizon cumulative relapse after a first depressive episode ^26^, short-term remission after acute treatment ^28,29^, and episode persistence over longer follow-up ^30^. For each domain, we distinguished calibration targets from benchmark comparisons used to evaluate model behavior. Full details are provided in the Supplement.

### Sensitivity Analyses

Parameter uncertainty was evaluated using probabilistic sensitivity analysis; reported 95% uncertainty intervals represent the 2.5th and 97.5th percentiles across 1000 parameter draws, with full distributional assumptions and simulation specifications provided in the Supplement.

We also conducted univariate sensitivity analyses varying key structural and design assumptions: chronic-relapse preventability with maintenance treatment, analytic time horizon, and monitoring frequency. For each scenario, we assessed whether strategy preference changed across risk strata and preference thresholds; scenario specifications are provided in the Supplement.

## Results

### Verifications and Validations

We assessed whether the model reproduced the clinical patterns it is intended to represent by comparing model projections with published clinical targets (Figure 2). Overall, the model tracked these benchmarks closely, reproducing the key features needed for the strategy comparisons: early post-remission relapse risk that attenuated over time, separation between maintenance and discontinuation arms, lower-risk cumulative relapse over multiple years, and persistence patterns consistent with episodic and chronic illness. For example, although calibration used relapse data only through 2 years after remission, the model generated a 32.0% 5-year cumulative relapse risk in the lower-risk group, compared with 31.5% (95% CI, 25.7%–37.3%) in a long-term cohort study of relapse following remission of a first lifetime depressive episode ^26^. Point-by-point comparisons of target and model values are shown in Figure 2 and Supplemental Table S2.

**Figure 2.**
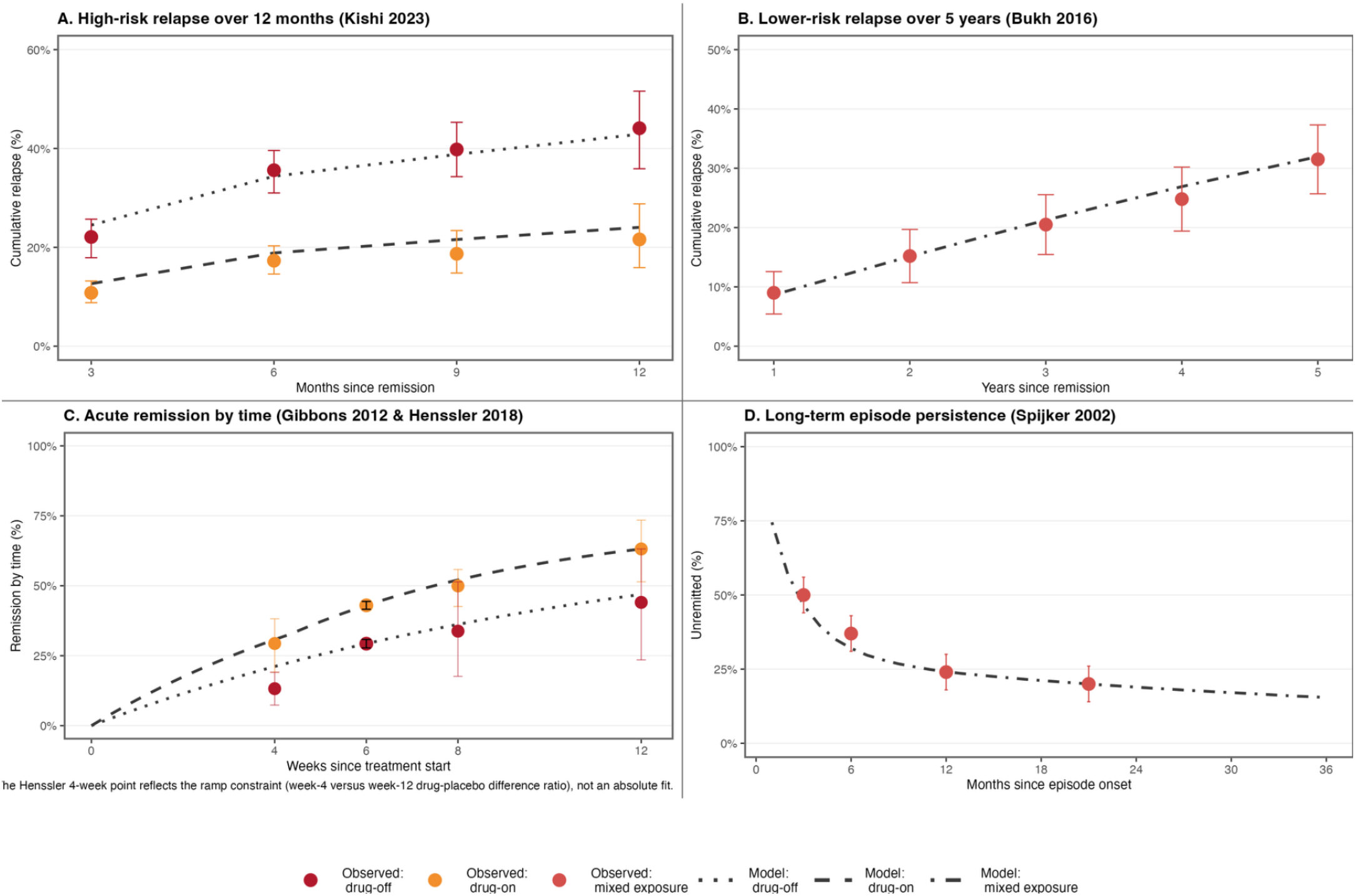
Model replicates external relapse, remission, and persistence benchmarks. Model projections (dashed lines) compared with published benchmarks (points with reported 95% CIs from the source studies) across four domains: (A) cumulative relapse over 12 months in high-risk patients from maintenance-discontinuation trials, (B) cumulative relapse over 5 years in lower-risk patients after a first lifetime depressive episode, (C) acute remission rates by time since treatment start, and (D) long-term episode persistence. Full details are provided in the Supplement.

### Base Case

In the base case, continuous maintenance resulted in fewer depression-months but substantially more medication exposure compared with active monitoring. The magnitude of this tradeoff varied markedly by baseline risk (Table 2). In the Very low group (calibrated to patients 8 months after remission from a first lifetime mild depressive episode), continuous maintenance required 11.8 medication-years per depression-month averted. The corresponding values for the Low, Moderate, and High groups were 5.1, 2.8, and 1.5 medication-years per depression-month averted, respectively.

**Table 2.**
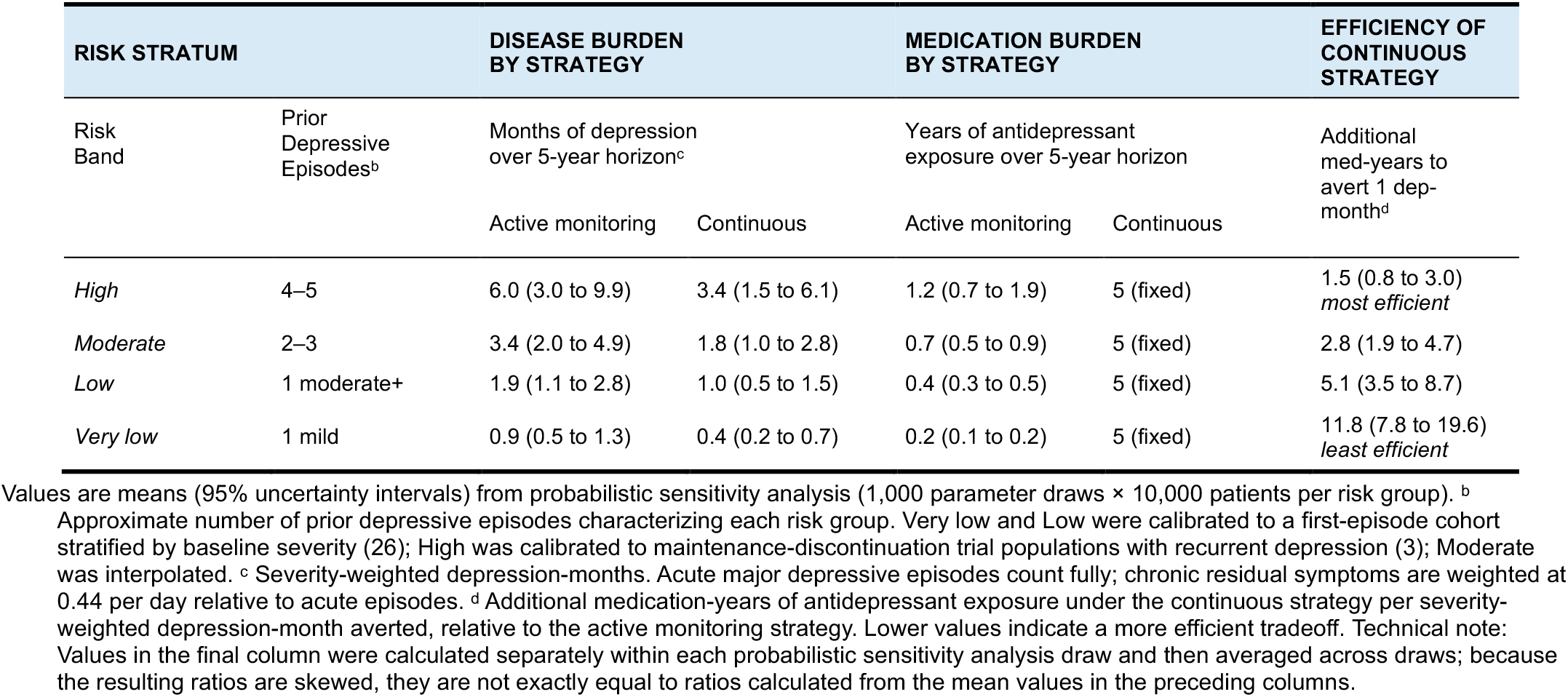
Base-Case Outcomes by Risk Group.

| RISK STRATUM |  | DISEASE BURDEN BY STRATEGY |  | MEDICATION BURDEN BY STRATEGY |  | EFFICIENCY OF CONTINUOUS STRATEGY |
| --- | --- | --- | --- | --- | --- | --- |
| Risk Band | Prior Depressive Episodes<sup>b</sup> | Months of depression over 5-year horizon<sup>c</sup> |  | Years of antidepressant exposure over 5-year horizon |  | Additional med-years to avert 1 dep-month<sup>d</sup> |
|  |  | Active monitoring | Continuous | Active monitoring | Continuous |  |
| *High* | 4–5 | 6.0 (3.0 to 9.9) | 3.4 (1.5 to 6.1) | 1.2 (0.7 to 1.9) | 5 (fixed) | 1.5 (0.8 to 3.0)<br>*most efficient* |
| *Moderate* | 2–3 | 3.4 (2.0 to 4.9) | 1.8 (1.0 to 2.8) | 0.7 (0.5 to 0.9) | 5 (fixed) | 2.8 (1.9 to 4.7) |
| *Low* | 1 moderate+ | 1.9 (1.1 to 2.8) | 1.0 (0.5 to 1.5) | 0.4 (0.3 to 0.5) | 5 (fixed) | 5.1 (3.5 to 8.7) |
| *Very low* | 1 mild | 0.9 (0.5 to 1.3) | 0.4 (0.2 to 0.7) | 0.2 (0.1 to 0.2) | 5 (fixed) | 11.8 (7.8 to 19.6)<br>*least efficient* |

Values are means (95% uncertainty intervals) from probabilistic sensitivity analysis (1,000 parameter draws × 10,000 patients per risk group). <sup>b</sup> Approximate number of prior depressive episodes characterizing each risk group. Very low and Low were calibrated to a first-episode cohort stratified by baseline severity (26); High was calibrated to maintenance-discontinuation trial populations with recurrent depression (3); Moderate was interpolated. <sup>c</sup> Severity-weighted depression-months. Acute major depressive episodes count fully; chronic residual symptoms are weighted at 0.44 per day relative to acute episodes. <sup>d</sup> Additional medication-years of antidepressant exposure under the continuous strategy per severity-weighted depression-month averted, relative to the active monitoring strategy. Lower values indicate a more efficient tradeoff. Technical note: Values in the final column were calculated separately within each probabilistic sensitivity analysis draw and then averaged across draws; because the resulting ratios are skewed, they are not exactly equal to ratios calculated from the mean values in the preceding columns.

In a decomposition of these outcomes, the additional depression burden under active monitoring was attributable primarily to episodes that were not prevented (82.3% of additional days depressed, averaged across risk groups), with smaller contributions from longer episode duration (9.0%) and their interaction (8.7%). Chronic residual symptoms accounted for the majority of depression burden under both strategies (63.8% under continuous maintenance; 53.1% under active monitoring).

### Incremental Net Benefit Analysis

Figure 3A presents the net benefit of continuous maintenance versus active monitoring, expressed in depression-months averted over 5 years, across varying preference thresholds for antidepressant exposure. The break-even threshold for each risk group equals its medication-years per depression-month averted from Table 2. Results are summarized at three illustrative thresholds (1, 2, and 3 medication-years per depression-month averted).

**Figure 3.**
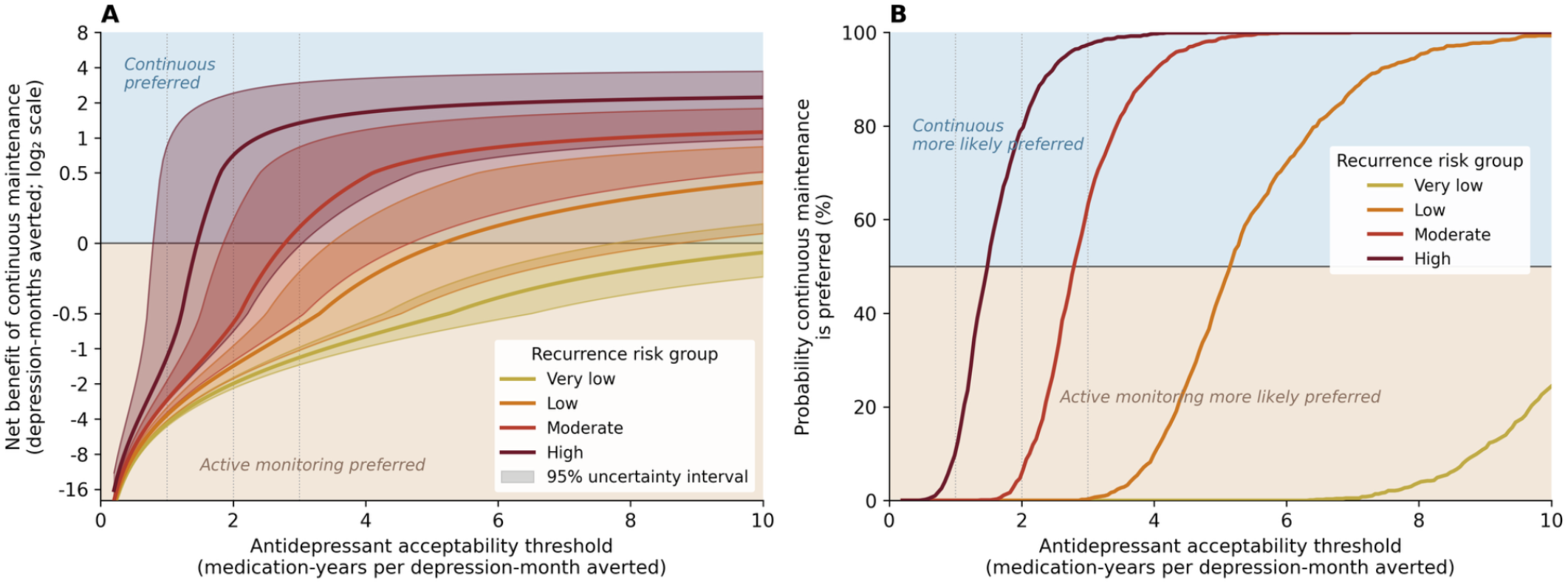
Net benefit and decision certainty for continuous maintenance versus active monitoring by preference threshold for antidepressant exposure. (A) Net benefit expressed as depression-months averted (log_2_ scale) over 5 years. Blue shading indicates the region where continuous maintenance is preferred; warm shading indicates where active monitoring is preferred. Solid lines show mean net benefit across probabilistic sensitivity analysis draws; shaded bands show 95% uncertainty intervals reflecting between-draw parameter uncertainty. (B) Probability that continuous maintenance is the preferred strategy at each preference threshold for antidepressant exposure. Vertical dotted lines in both panels mark the three illustrative preference thresholds (1, 2, and 3 medication-years per depression-month averted).

At the lowest preference threshold (1 medication-year per depression-month averted), continuous maintenance produced net harm across all risk groups. At the intermediate threshold (2 medication-years per depression-month averted), maintenance showed net benefit only in the High group. At the highest threshold (3 medication-years per depression-month averted), maintenance showed net benefit in the High group and crossed the break-even point in the Moderate group, although uncertainty in the Moderate group overlapped zero. Thus, even when patients strongly prioritized avoiding depression, expected gains from maintenance concentrated in the Moderate and High groups.

In probabilistic preference curves (Figure 3B), maintenance was increasingly preferred as relapse risk and willingness to accept medication exposure increased.

### Univariate Sensitivity Analyses

To evaluate robustness, we examined 6 univariate sensitivity scenarios varying clinical monitoring frequency, analytic time horizon, and the preventability of chronic depression with maintenance treatment (Tables S3). We classified the base-case preferred strategy for each combination of risk group and preference threshold, then assessed how often these classifications changed across scenarios. Overall, 68 of 72 (94%) cell-level classifications were unchanged. The few shifts occurred near break-even regions and moved in expected directions: modeling chronic depression as more preventable and extending the time horizon favored continuous maintenance more strongly, whereas more frequent monitoring somewhat improved the favorability of active monitoring. The largest shifts arose in the no chronic-prevention-benefit scenario, in which at 2 medication-years per depression-month averted no group favored maintenance and at 3 only the High-risk group favored maintenance. Overall, these findings suggest that the model’s conclusions are robust and driven primarily by patient preferences and baseline relapse risk rather than by plausible variation in other assumptions.

## Discussion

In this decision-analytic model of adults who had completed an 8-month continuation phase after remission from major depressive disorder, continuous maintenance reduced depression burden over 5 years but required substantially more antidepressant exposure than active monitoring. The efficiency of this tradeoff increased markedly with baseline relapse risk: averting 1 month of depression required roughly 1.5 additional years of antidepressant exposure in the highest-risk group, compared with more than a decade in the lowest-risk group.

These findings are broadly consistent with current guideline logic ^12–15^: maintenance becomes more compelling as relapse risk rises, but the preferred strategy also depends on how much medication exposure a patient is willing to accept to avert future depression.

The model extends guideline reasoning most consequentially for the Moderate-risk group—conceptually, the large group between first-episode and clearly recurrent illness, approximated here by patients with 2 to 3 prior episodes who are several months past remission. For these patients, the break-even preference threshold was approximately 2.8 medication-years per depression-month averted: continuous maintenance is likely to be value-concordant only if patients are at least moderately accepting of long-term antidepressant use, whereas active monitoring may be preferred for those more reluctant to continue medication.

The framework also adds nuance at the extremes: maintenance may remain value-concordant for Low-risk patients willing to accept about 5.1 medication-years per depression-month averted, while active monitoring may remain preferred for High-risk patients unwilling to accept more than 1.5.

To make this logic concrete, consider a 26-year-old woman with 2 years remaining in graduate student several months past remission from a first lifetime moderately severe depressive episode (Low-risk group). After discussing the tradeoffs, she might say: “I’d continue antidepressants through graduate school to prevent another month of depression, but not through my twenties unless they prevented multiple months.” Her implied preference threshold is at least 2 and at most 4 medication-years per depression-month averted. Because this falls below the Low-risk break-even threshold of about 5, active monitoring would be favored. If, a decade later, she had experienced several relapses and resembled the High-risk profile, the same preference threshold would favor continuous maintenance.

These thresholds convert broad guideline recommendations, including general calls for shared decision-making, into explicit patient-facing tradeoffs: additional time taking antidepressants while well versus time spent depressed ^17^.

The relapse-risk strata used here should be interpreted as clinically anchored benchmarks rather than individualized prediction categories. Patients may not map cleanly onto a single stratum, and relapse risk may be modified by residual symptoms, comorbidity, suicidality, substance use, social stressors, age, treatment history, and other factors not represented directly in the model. Future work could replace these strata with individualized relapse-risk estimates from validated multivariable prediction models. The purpose of the present approach was to make the scale of the maintenance-versus-monitoring tradeoff visible across recognizable regions of relapse risk, not to provide deterministic bedside predictions. Even when a patient’s exact risk category is uncertain, recognizing that the patient is closer to the Moderate or High benchmark than to the Low benchmark may be sufficient to inform shared decision-making when considered alongside that patient’s preference threshold for antidepressant exposure.

The large contribution of chronic residual symptoms also suggests that future decision-support models should distinguish risk of relapse from risk of prolonged or chronic depressive course. A relapse that resolves quickly and a relapse that leaves persistent residual symptoms have different consequences for the maintenance decision, even if both count as relapse events in trial-based evidence.

This study has several limitations. First, we did not model withdrawal symptoms, rebound effects, tolerance, potentiation, or the challenges patients can have in restarting or tapering antidepressants. Second, the preference thresholds in this framework are illustrative quantities, not empirically measured patient preferences; future work should characterize their distribution and stability in clinical populations. Third, the model isolates the maintenance-versus-monitoring tradeoff for antidepressant pharmacotherapy and is not intended as a comprehensive model of long-term depression care. Factors not represented include alternative or adjunctive treatments such as psychotherapy, exercise therapy, and neuromodulation, as well as comorbidity, suicidality, substance use, social stressors, life events, medication dose, adherence,functioning, and quality of life. Fourth, evidence remains contested regarding the magnitude and interpretation of antidepressant relapse-prevention effects ^32–34^. As such, the modeling framework provided herein should not be used to replace clinical judgment.

Future work should test decision support tools that combine individualized relapse-risk estimates with patient preferences to guide maintenance decisions in real-world care, including testing the feasibility of preference elicitation and stability of expressed preferences. If future research can better predict periods of highest relapse risk, treatment could be intensified selectively—preserving much of the benefit of maintenance while reducing unnecessary exposure for patients unlikely to benefit.

In this decision-analytic study, continuous maintenance reduced depression burden compared with active monitoring, but the efficiency of the tradeoff between additional antidepressant exposure and depression-months averted varied substantially by relapse risk. This work provides a quantitative framework for antidepressant maintenance decisions, showing that the value of continuous maintenance depends strongly on baseline relapse risk and patient preferences regarding long-term antidepressant exposure.

## Data Availability

No new individual-level human participant data were generated or analyzed. All model inputs were aggregate estimates obtained from the cited publications and supplementary materials, and the key outputs of the model are reported in the manuscript and Supplement. The analytic code is available from the corresponding author upon reasonable request.

## Article Information

### Author Contributions

Dr Meyerson had full access to all model inputs, analytic code, and model outputs and takes responsibility for the integrity of the data, accuracy of the analysis, and fidelity of the manuscript to the reported results.

Concept and design: Meyerson.

Acquisition, analysis, or interpretation of data: All authors

Drafting of the manuscript: Meyerson.

Critical revision of the manuscript for important intellectual content: All authors

Statistical analysis: Meyerson.

Obtained funding: NA

Administrative, technical, or material support: Smoller.

Supervision: Cai, Smoller.

### Data Sharing Statement

This decision-analytic modeling study used model inputs derived from published aggregate data. No individual participant data were analyzed. The underlying published data sources are cited in the manuscript and Supplement.

### Use of AI-assisted technologies

OpenAI ChatGPT GPT-5 Thinking-series models (OpenAI, San Francisco, California) and Anthropic Claude Opus/Sonnet 4-series models (Anthropic, San Francisco, California) were used intermittently from October 2025 through May 2026 in an assistive role during study development and manuscript preparation, including code refactoring and editing, discussion of analytic implementation details, and drafting, revising, and editing portions of the manuscript text. The study concept, design, core analytic decisions, original code framework, results interpretation, and final manuscript content were determined and verified by the authors. All AI-assisted output was reviewed, revised as needed, and checked by the authors, who take full responsibility for the accuracy and integrity of the work.

## References

1. Hasin DS, Sarvet AL, Meyers JL, et al. Epidemiology of adult DSM-5 major depressive disorder and its specifiers in the United States. JAMA Psychiatry. 2018;75(4):336–346. doi:10.1001/jamapsychiatry.2017.4602

2. Monroe SM, Harkness KL. Major depression and its recurrences: life course matters. Annu Rev Clin Psychol. 2022;18:329–357. doi:10.1146/annurev-clinpsy-072220-021440

3. Kishi T, Sakuma K, Hatano M, et al. Relapse and its modifiers in major depressive disorder after antidepressant discontinuation: meta-analysis and meta-regression. Mol Psychiatry. 2023;28(3):974–976. doi:10.1038/s41380-022-01920-0

4. Kato M, Hori H, Inoue T, et al. Discontinuation of antidepressants after remission with antidepressant medication in major depressive disorder: a systematic review and meta-analysis. Mol Psychiatry. 2021;26(1):118–133. doi:10.1038/s41380-020-0843-0

5. Cartwright C, Gibson K, Read J, Cowan O, Dehar T. Long-term antidepressant use: patient perspectives of benefits and adverse effects. Patient Prefer Adherence. 2016;10:1401–1407. doi:10.2147/PPA.S110632

6. Reichenpfader U, Gartlehner G, Morgan LC, et al. Sexual dysfunction associated with second-generation antidepressants in patients with major depressive disorder: results from a systematic review with network meta-analysis. Drug Saf. 2014;37(1):19–31. doi:10.1007/s40264-013-0129-4

7. Gafoor R, Booth HP, Gulliford MC. Antidepressant utilisation and incidence of weight gain during 10 years’ follow-up: population based cohort study. BMJ. 2018;361:k1951. doi:10.1136/bmj.k1951

8. Zaccoletti D, Mosconi C, Gastaldon C, et al. Comparison of antidepressant deprescribing strategies in individuals with clinically remitted depression: a systematic review and network meta-analysis. Lancet Psychiatry. 2026;13(1):24–36. doi:10.1016/S2215-0366(25)00330-X

9. Zhu JM, Huntington A, Haeder S, Wolk C, McConnell KJ. Insurance acceptance and cash pay rates for psychotherapy in the US. Health Aff Sch. 2024;2(9):qxae110. doi:10.1093/haschl/qxae110

10. Mojtabai R, Olfson M, Sampson NA, et al. Barriers to mental health treatment: results from the National Comorbidity Survey Replication. Psychol Med. 2011;41(8):1751–1761. doi:10.1017/S0033291710002291

11. Hockenberry JM, Joski P, Yarbrough C, Druss BG. Trends in treatment and spending for patients receiving outpatient treatment of depression in the United States, 1998-2015. JAMA Psychiatry. 2019;76(8):810–817. doi:10.1001/jamapsychiatry.2019.0633

12. National Institute for Health and Care Excellence. Depression in adults: treatment and management. NICE; 2022. Accessed April 1, 2024. https://www-nice-org-uk.proxy.lib.duke.edu/guidance/ng222/chapter/Recommendations#further-line-treatment

13. McQuaid JR, Buelt A, Capaldi V, et al. The management of major depressive disorder: synopsis of the 2022 U.S. Department of Veterans Affairs and U.S. Department of Defense clinical practice guideline. Ann Intern Med. 2022;175(10):1440–1451. doi:10.7326/M22-1603

14. Lam RW, Kennedy SH, Adams C, et al. Canadian Network for Mood and Anxiety Treatments (CANMAT) 2023 update on clinical guidelines for management of major depressive disorder in adults. Can J Psychiatry. 2024;69(9):641–687. doi:10.1177/07067437241245384

15. American Psychiatric Association. Practice Guideline for the Treatment of Patients With Major Depressive Disorder. 3rd ed. American Psychiatric Association; 2010.

16. Wang JL, Patten S, Sareen J, Bolton J, Schmitz N, MacQueen G. Development and validation of a prediction algorithm for use by health professionals in prediction of recurrence of major depression. Depress Anxiety. 2014;31(5):451–457. doi:10.1002/da.22215

17. Stacey D, Lewis KB, Smith M, et al. Decision aids for people facing health treatment or screening decisions. Cochrane Database Syst Rev. 2024;1(1):CD001431. doi:10.1002/14651858.CD001431.pub6

18. Hu Y, Xue H, Ni X, Guo Z, Fan L, Du W. Association between duration of antidepressant treatment for major depressive disorder and relapse rate after discontinuation: a meta-analysis. Psychiatry Res. 2024;337:115926. doi:10.1016/j.psychres.2024.115926

19. Judd LL, Akiskal HS, Maser JD, et al. A prospective 12-year study of subsyndromal and syndromal depressive symptoms in unipolar major depressive disorders. Arch Gen Psychiatry. 1998;55(8):694–700. doi:10.1001/archpsyc.55.8.694

20. Lokkerbol J, Wijnen B, Ruhe HG, et al. Design of a health-economic Markov model to assess cost-effectiveness and budget impact of the prevention and treatment of depressive disorder. Expert Rev Pharmacoecon Outcomes Res. 2021;21(5):1031–1042. doi:10.1080/14737167.2021.1844566

21. Sobocki P, Ekman M, Ovanfors A, Khandker R, Jönsson B. The cost-utility of maintenance treatment with venlafaxine in patients with recurrent major depressive disorder. Int J Clin Pract. 2008;62(4):623–632. doi:10.1111/j.1742-1241.2008.01711.x

22. Nuijten MJC. Assessment of clinical guidelines for continuation treatment in major depression. Value Health. 2001;4(4):281–294. doi:10.1046/j.1524-4733.2001.44053.x

23. Meyerson WU, Ross EL, Kennedy CJ, et al. Aripiprazole or bupropion augmentation versus switching to bupropion in treatment-resistant depression: a risk-benefit analysis. J Clin Psychiatry. 2025;86(4):25m15863. doi:10.4088/JCP.25m15863

24. Gold MR. Cost-Effectiveness in Health and Medicine. Oxford University Press; 1996.

25. Husereau D, Drummond M, Augustovski F, et al. Consolidated Health Economic Evaluation Reporting Standards 2022 (CHEERS 2022) statement: updated reporting guidance for health economic evaluations. MDM Policy Pract. 2022;7(1):23814683211061097. doi:10.1177/23814683211061097

26. Bukh JD, Andersen PK, Kessing LV. Rates and predictors of remission, recurrence and conversion to bipolar disorder after the first lifetime episode of depression: a prospective 5-year follow-up study. Psychol Med. 2016;46(6):1151–1161. doi:10.1017/S0033291715002676

27. Zhou D, Lv Z, Shi L, et al. Effects of antidepressant medicines on preventing relapse of unipolar depression: a pooled analysis of parametric survival curves. Psychol Med. 2022;52(1):48–56. doi:10.1017/S0033291720001610

28. Gibbons RD, Hur K, Brown CH, Davis JM, Mann JJ. Benefits from antidepressants: synthesis of 6-week patient-level outcomes from double-blind placebo-controlled randomized trials of fluoxetine and venlafaxine. Arch Gen Psychiatry. 2012;69(6):572–579. doi:10.1001/archgenpsychiatry.2011.2044

29. Henssler J, Kurschus M, Franklin J, Bschor T, Baethge C. Trajectories of acute antidepressant efficacy: how long to wait for response? A systematic review and meta-analysis of long-term, placebo-controlled acute treatment trials. J Clin Psychiatry. 2018;79(3):17r11470. doi:10.4088/jcp.17r11470

30. Spijker J, de Graaf R, Bijl RV, Beekman ATF, Ormel J, Nolen WA. Duration of major depressive episodes in the general population: results from the Netherlands Mental Health Survey and Incidence Study. Br J Psychiatry. 2002;181(3):208–213. doi:10.1192/bjp.181.3.208

31. Spijker J, Bijl RV, de Graaf R, Nolen WA. Care utilization and outcome of DSM-III-R major depression in the general population: results from the Netherlands Mental Health Survey and Incidence Study. Acta Psychiatr Scand. 2001;104(1):19–24. doi:10.1034/j.1600-0447.2001.00363.x

32. Ghaemi SN, Selker HP. Maintenance efficacy designs in psychiatry: randomized discontinuation trials—enriched but not better. J Clin Transl Sci. 2017;1(3):198–204. doi:10.1017/cts.2017.2

33. Hengartner MP, Plöderl M. Prophylactic effects or withdrawal reactions? An analysis of time-to-event data from antidepressant relapse prevention trials submitted to the FDA. Ther Adv Psychopharmacol. 2021;11:20451253211032051. doi:10.1177/20451253211032051

34. Horowitz MA, Taylor D. Distinguishing relapse from antidepressant withdrawal: clinical practice and antidepressant discontinuation studies. BJPsych Adv. 2022;28(5):297–311. doi:10.1192/bja.2021.62

35. Masdrakis VG, Markianos M, Baldwin DS. Apathy associated with antidepressant drugs: a systematic review. Acta Neuropsychiatr. 2023;35(4):189–204

