## Supplementary Appendix 1 for "Antidepressant Maintenance Versus Active Monitoring After Depression Remission: A Decision Analysis Stratified by Relapse Risk and Patient Preferences"

Supplementary Materials

### I. Parameter Derivations

The parameter derivations below refer to the health state transition model shown in Supplemental Figure S1.

Supplemental Figure S1. Health state transition model schematic.

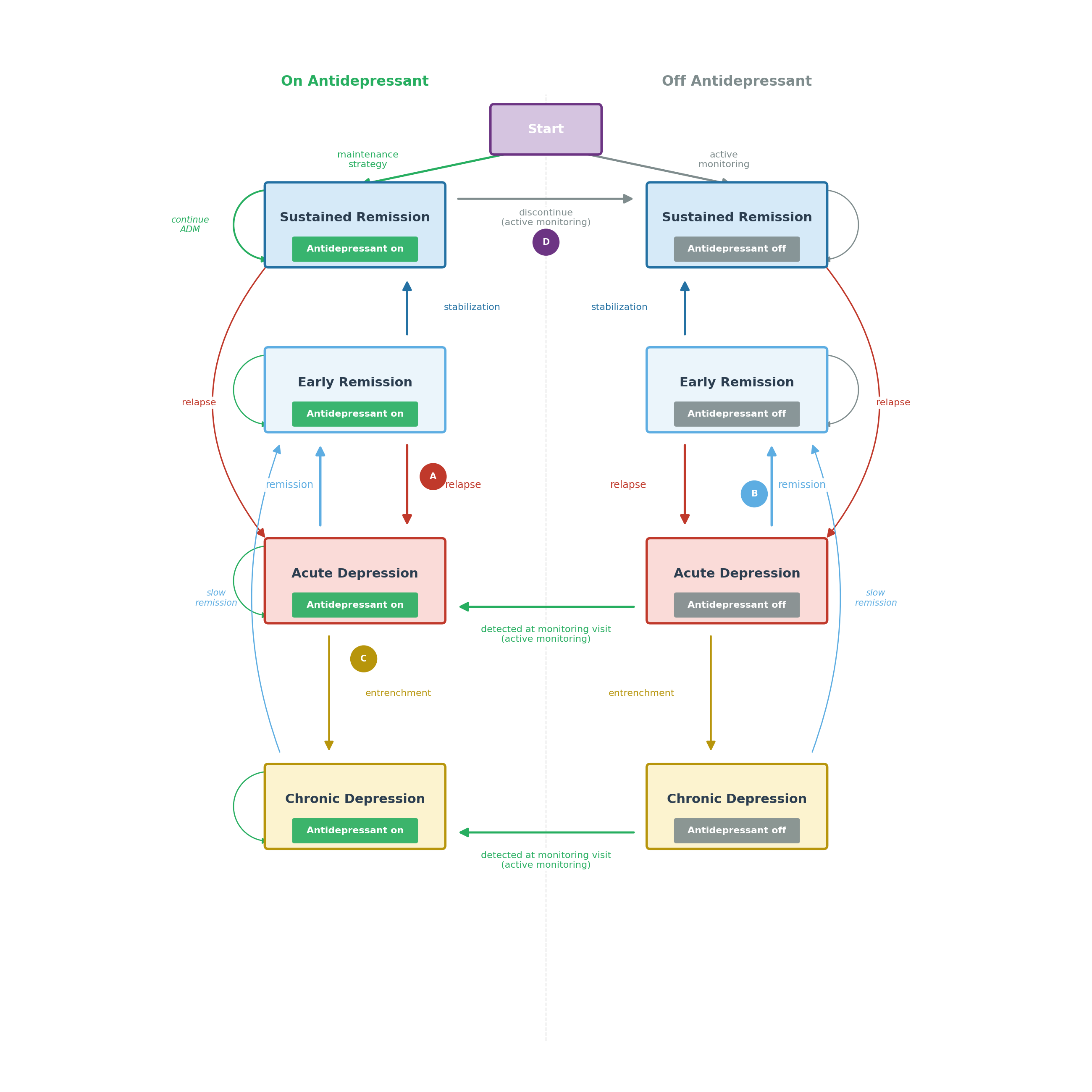

Supplemental Figure S1: States are shown separately for on- versus off-antidepressant treatment. Arrows indicate strategy assignment at model entry, remission, relapse, stabilization, chronic-course entrenchment, and monitoring-triggered transitions back to on-antidepressant states under active monitoring; self-loops indicate persistence within states from one monthly cycle to the next.

#### A. Relapse Parameters

##### A0) Overview

To parameterize relapse for a decision-focused simulation, we reviewed the maintenance/discontinuation and longitudinal-course literature with the practical goal of identifying sources that (a) report arm-specific relapse over time under randomized discontinuation versus maintenance, (b) provide longer-horizon follow-up beyond the first year, and (c) inform relapse rates in both lower-risk and higher-risk populations. The sources we relied on most for relapse parameter derivation were Kishi et al. (2023), Zhou et al. (2020), and Bukh et al. (2016), because together they capture short-horizon maintenance/discontinuation effects, time-patterning over the first year, and longer-horizon recurrence in a lower-risk cohort (3,27,28).

Kishi et al. (2023) is a systematic review and meta-analysis of maintenance/discontinuation randomized trials. It summarizes relapse patterns under maintenance and discontinuation in a population with highly recurrent depression, with an average of approximately 4 to 5 lifetime episodes according to its supplemental tables (3). Several empirical patterns were evident within the first 12 months of follow-up, whereas later time points were less consistently supported across trials. These included (i) substantial cumulative relapse under discontinuation, with 44.1% relapse in the placebo arm by 12 months; (ii) front-loaded relapse accumulation, with a higher-risk early period after randomization followed by a flatter, more nearly linear accumulation later in follow-up; (iii) a robust preventive effect of antidepressant maintenance; and (iv) similar relative separation between maintenance and placebo across the best-supported follow-up time points (3). As a matter of time indexing, Kishi et al. (2023) defined T=0 at randomization, which we estimated to occur about 2 months after clinical remission based on the distribution of continuation periods reported in the largely overlapping meta-analysis by Zhou et al. (2020) (27).

Bukh et al. (2016), a first-episode observational cohort with up to 5 years of follow-up, informed the lower-risk strata and longer-horizon behavior in three ways: (v) cumulative recurrence was substantially lower than in the highly recurrent trial populations synthesized by Kishi et al. (2023); (vi) recurrence was still lower in the subgroup with mild index episodes; and (vii) after the early post-remission period, recurrence accumulated at an approximately steady background rate over extended follow-up, consistent with roughly linear accumulation from years 1 through 5 (26).

Together, these observations motivated a small set of explicit modeling assumptions used to translate these sources into a monthly relapse process: (A) relapse risk is represented as a late-period (“steady-state”) monthly hazard with an early post-remission elevation that attenuates over the first months after discontinuation or randomization; (B) the placebo/discontinuation arm in Kishi et al. (2023) defines the high-risk anchor for off-treatment relapse risk in a highly recurrent population; (C) antidepressant maintenance is modeled as a constant on-treatment hazard ratio; (D) the two lowest-risk strata are anchored to Bukh et al. (2016), using its mild versus non-mild first-episode groups to define the Very low- and Low-risk strata; and (E) an intermediate Moderate-risk stratum, notionally corresponding to an intermediate recurrence history such as 2 to 3 lifetime episodes, is defined by interpolation between the low-risk and high-risk anchors.

The full set of model parameters, including relapse, remission, chronic-course, and model-structure inputs, is presented in Table S1 below.

**Table S1. Base-case model parameters, uncertainty ranges, and data sources**

| **Parameter** | **Base case** | **Units** | **Range / Scenarios** | **Source** |
| --- | --- | --- | --- | --- |
| ***Relapse risk by patient group*** |  |  |  |  |
| Very low risk (first episode, mild severity) | 0.0033 | prob/month | 0.0023 to 0.0046 | Bukh et al. (2016) |
| Low risk (first episode, moderate+ severity) | 0.0072 | prob/month | 0.005 to 0.010 | Bukh et al. (2016) |
| Moderate risk (interpolated) | 0.013 | prob/month | 0.009 to 0.018 | Interpolated between Low and High groups |
| High risk (4–5 prior episodes) | 0.023 | prob/month | 0.013 to 0.040 | Kishi et al. (2023) |
| ***Relapse risk modifiers*** |  |  |  |  |
| Observed composite hazard ratio for relapse prevention | 0.470 | HR | 0.420 to 0.530 | Zhou et al. (2020) |
| Decomposed non-chronic relapse HR | 0.413 | HR | Derived from composite | See Section A5 |
| Implied chronic relapse HR | 0.643 | HR | Derived from composite | See Section A5 |
| Chronic relapse effect fraction | 0.50 | Proportion | 0, 0.5, 1.0 | Assumption |
| Hazard multiplier months 1–5 | 4.00856 | multiplier | fixed | Fit to Kishi et al. (2023) |
| Hazard multiplier months 6–8 | 2.00214 | multiplier | fixed | Fit to Kishi et al. (2023) |
| ***Remission*** |  |  |  |  |
| Remission HR, on- vs off-ADM | 2.08372 | HR | 1.86 to 6.12 | Gibbons et al. (2012) |
| Monthly remission prob (off ADM; non-chronic) | 0.314 | prob/month | 0.272 to 0.812 | Gibbons et al. (2012) |
| Treatment effect ramp (month 1) | 0.636 | fraction | 0.519 to 0.751 | Henssler et al. (2018) |
| ***Chronic depression*** |  |  |  |  |
| Base probability chronic course | 0.292 | proportion | 0.185 to 0.729 | Spijker et al. (2002) |
| Chronic residual symptoms burden weight | 0.445 | burden/day | 0.394 to 0.495 | Judd et al. (1998) |
| Monthly remission prob (off ADM; chronic) | 0.014 | prob/month | 0.000 to 0.052 | Spijker et al. (2002) |
| Chronic relapse prevention fraction | 0.500 | fraction | 0, 0.5, 1 | Assumption |
| ***Model structure*** |  |  |  |  |
| Continuation phase before taper, months | 8 | months | fixed | CANMAT 2023, VA/DoD 2022 |
| Monitoring interval (active monitoring) | 3 | months | 1, 3, 6 months | Clinical practice |
| Time horizon, years | 5 | years | 3, 5, 10 years | Bukh et al. (2016) follow-up duration |

Abbreviations: ADM, antidepressant medication; HR, hazard ratio; RR, relative risk. Uncertainty ranges are 95% CIs used in probabilistic sensitivity analysis. Scenario parameters are varied in univariate sensitivity analyses.

##### A1) High-risk stratum: baseline late-period off-treatment relapse hazard.

***In brief:*** *We isolated the 6-to-12-month segment of the Kishi et al. (2023) placebo arm—where relapse accumulation is approximately linear—and solved for the constant monthly hazard that reproduces it (h ≈ 0.023/month).*

**In detail:** The high-risk stratum's relapse risk was anchored to Kishi et al. (2023). Specifically, the "steady state" off-antidepressant relapse hazard in the high-risk stratum was derived from the placebo arm of Kishi et al. (2023) by isolating relapse accumulation over the later portion of follow-up and solving for the constant monthly hazard that reproduces the increase in cumulative relapse from 6 to 12 months.

Let R6 and R12 denote the digitized placebo-arm cumulative relapse at 6 and 12 months post-randomization. Under a constant hazard over this six-month interval, survival from month 6 to month 12 satisfies (1 − R12)/(1 − R6) = exp(−6h), yielding h = −log[(1 − R12)/(1 − R6)]/6 for the late-period monthly hazard.

$$\frac{\left( 1-R_{12} \right)}{\left( 1-R_{6} \right)}=exp\left( -6h \right)$$

This closed-form expression was used to estimate the late-period monthly hazard h.

As a matter of accounting, we apply this hazard starting at post-remission month 9 (the month that starts at 8 and ends at 9) and onwards, after accounting for the fact that months are indexed in Kishi et al. (2023) from time of randomization rather than from time of remission.

Uncertainty was obtained by sampling the Kishi et al. (2023) placebo cumulative relapse targets at 6 and 12 months from their reported 95% confidence intervals and recomputing the implied late-period hazard h across draws, yielding a distribution for the high-risk late-period off-treatment hazard.

##### A2) Very low- and Low-risk strata: baseline late-period off-treatment relapse hazards.

***In brief:*** *We calibrated two low-risk strata to Bukh et al. 2016’s 2-year recurrence after a first episode, using its mild/moderate-severe severity split (HR = 2.2) to separate Very low- from Low-risk, and accounting for assumed guideline-concordant continuation treatment in the cohort (h_vlow ≈ 0.0033/month; h_low ≈ 0.0072/month).*

**In detail:** The two lowest-risk strata were anchored to Bukh et al. (2016), an observational cohort of patients remitted from a first lifetime depressive episode with follow-up out to five years. We used Bukh et al. (2016)’s severity split (mild vs moderate-to-severe) to define two low-risk strata.

We mapped Bukh et al. (2016)’s mild subgroup to the Very low-risk stratum and the moderate-to-severe subgroup to the Low-risk stratum. Note that in Bukh et al. (2016), “moderate-to-severe” refers to baseline episode severity among first-episode patients (predominantly moderate severity); these patients had only one lifetime episode and thus represent a low-recurrence-risk group despite having at least moderate episode severity. In Bukh et al. (2016), the proportion of episodes that were mild was 0.243, the proportion that were moderate-to-severe was 0.757, and the reported recurrence hazard ratio comparing moderate-to-severe vs mild was 2.2. We therefore parameterized the Low-risk late-period hazard as proportional to the Very low-risk hazard: h_low = 2.2 × h_vlow.

To identify h_vlow, we calibrated to Bukh et al. (2016)’s overall 2-year cumulative recurrence (15.2%) by requiring the model’s pooled 2-year recurrence—combining mild and moderate-to-severe using Bukh et al. (2016)’s observed proportions—to match this target. We used the same time-since-remission hazard multipliers w_m (Section I.A4), with w_m = 1 beyond the first-year shaping window.

Because Bukh et al. (2016) does not standardize antidepressant exposure after remission, we made an explicit accounting assumption to interpret its recurrence in terms of off-treatment hazards: we assumed guideline-concordant continuation treatment for 8 months after remission followed by discontinuation. Let e_m denote the medication exposure factor implied by this assumption (e_m = HR_maintenance during continuation months; e_m = 1 after discontinuation). The calibration solves for h_vlow such that the weighted 2-year recurrence from the Very low and Low strata matches the reported 15.2% recurrence target.

Here, m = 3..26 indexes months starting after the 2-month remission requirement in Bukh et al. (2016). Once h_vlow was solved from this calibration, we set h_low = 2.2 × h_vlow.

Uncertainty was propagated by sampling the Bukh et al. (2016) recurrence target from its reported uncertainty and re-solving for h_vlow across draws, yielding distributions for h_vlow and h_low.

##### A3) Moderate-risk stratum: baseline late-period off-treatment relapse hazard.

***In brief:*** *The Moderate-risk hazard was set to the geometric midpoint of the Low- and High-risk hazards on the log scale—an interpolation that avoids introducing an additional free parameter (h_mod ≈ 0.013/month).*

**In detail:** We defined the Moderate-risk stratum as an intermediate relapse-propensity group between the Low-risk first-episode anchor (non-mild severity, one lifetime episode; Section I.A2) and the highly recurrent trial-based anchor (Section I.A1). Notionally, this stratum represents patients with an intermediate recurrence history (e.g., two or three prior episodes), but no single source provided a sufficiently clean match to that population while also reporting the time-specific relapse quantities needed for our monthly model. We therefore defined the Moderate-risk late-period off-treatment hazard by interpolation between the two empirically anchored endpoints.

Specifically, we set the Moderate-risk late-period hazard to the geometric midpoint of the Low- and High-risk hazards (log-scale interpolation): h_mod = exp[(log h_low + log h_high)/2]. This yields a monotone, proportional mapping without introducing an additional free parameter.

Uncertainty for h_mod was obtained by propagating the uncertainty already assigned to the Low- and High-risk hazards. In each probabilistic draw, we take one plausible Low-risk hazard value and one plausible High-risk hazard value and then define the Moderate-risk hazard as the proportional midpoint between them.

##### A3a) Population context for relapse-risk strata

The relapse-risk strata were designed as clinically anchored benchmarks rather than formal population-prevalence categories. To provide context for how recognizable these strata may be in practice, we conducted a descriptive analysis of public-use National Comorbidity Survey Replication (NCS-R) data (35,36). We grouped adults with DSM-IV major depressive episode by reported lifetime number of depressive episodes, using categories corresponding approximately to the model’s recurrence-history anchors: 1 lifetime episode, 2 to 3 lifetime episodes, and 4 or more lifetime episodes. Survey-weighted estimates were calculated for adults with lifetime major depressive episode and for adults with 12-month major depressive episode. The lifetime denominator provides a person-level view of how commonly individuals ever fall into these episode-history groups, whereas the 12-month denominator better approximates the recent-depression population from which post-remission maintenance decisions may arise.

Among adults with lifetime DSM-IV major depressive episode, 28.7% reported 1 lifetime episode, 29.4% reported 2 to 3 episodes, and 41.9% reported 4 or more episodes. Among adults with 12-month DSM-IV major depressive episode, the corresponding proportions were 14.8%, 24.6%, and 60.6%. Because the survey episode-count categories do not distinguish first mild episodes from first moderate-to-severe episodes, the Very low versus Low first-episode split was contextualized using the first-episode cohort that informed the lower-risk relapse hazards. In that cohort, 24.3% of first-episode participants were classified as mild and 75.7% as moderate-to-severe (26).

Combining these sources suggests that, among adults with 12-month DSM-IV major depressive episode, the approximate contextual distribution corresponding to the model strata is 3.6% Very low, 11.2% Low, 24.6% Moderate, and 60.6% High. Using the lifetime DSM-IV major depressive episode denominator, the corresponding estimates are 7.0%, 21.7%, 29.4%, and 41.9%. These values should not be interpreted as definitive prevalence estimates for the modeled strata, because the model combines episode history, severity, and trial- or cohort-derived relapse hazards rather than applying a single survey-derived classification rule. They nevertheless support the clinical recognizability of the modeled anchors: first-episode, intermediate recurrent, and highly recurrent groups are all identifiable in nationally representative data, and recurrent illness accounts for a large share of adults with recent major depressive episode.

##### A4) Early-post-remission hazard shaping (months 1–8):

***In brief:*** *We fit a single free parameter—an early-post-remission hazard multiplier—to reproduce the front-loaded shape of the Kishi et al. (2023) placebo trajectory, with a log-linear stepdown to steady state by month 9 (w_early ≈ 4.0; w_mid ≈ 2.0).*

In detail: We modeled early post-remission relapse risk by multiplying each stratum’s late-period off-treatment hazard h_stratum by time-since-remission multipliers w_m, so that the monthly off-treatment hazard is h_stratum × w_m.

The w_m pattern was derived from digitized placebo-arm cumulative relapse in Kishi et al. (2023) at 3, 6, 9, and 12 months post-randomization. Because Kishi et al. (2023) indexes time from randomization whereas our model indexes time from remission, we shifted the Kishi et al. (2023) timepoints onto the remission clock using the same remission-to-randomization lag as in Section I.A1 (rounded to 2 months). Under our month indexing convention, “month 9” refers to the interval from post-remission month 8 to 9; accordingly, the late-period (“steady-state”) regime begins at month 9 on the remission-based clock, i.e., w_m = 1 for m ≥ 9.

We therefore used an early elevated level w_early for months 1–5 and a transition level w_mid for months 6–8 that bridges from w_early down to 1, with w_m = 1 from month 9 onward on the remission-based clock. Only one parameter was free: w_early. Given w_early, we set w_mid as the log-linear midpoint between w_early and 1 (equivalently, w_mid = sqrt[w_early]), and then chose w_early to best reproduce the shifted Kishi et al. (2023) placebo cumulative relapse trajectory.

We treated w_m as fixed (i.e., did not assign separate uncertainty) because these multipliers are highly correlated with the baseline late-period hazards; instead, we propagate uncertainty through the stratum-specific late hazards.

##### A5) On-treatment effect: maintenance vs discontinuation hazard ratio for relapse.

***In brief:*** *We took the maintenance-versus-discontinuation relapse hazard ratio from Zhou et al. (2020)’s pooled survival model at 6 months (HR ≈ 0.47, 95% CI 0.42–0.53). Because this was observed in a mixed chronic/non-chronic population, we decomposed it into a stronger non-chronic HR (≈0.41) and a weaker chronic HR (≈0.64). The observed composite (0.47) is used for calibration; the decomposed non-chronic HR is used in the simulation.*

**In detail:** Kishi et al. (2023) reports maintenance effects primarily as relative risks for cumulative relapse rather than a hazard ratio, whereas our simulation applies treatment effects on the hazard scale. We therefore obtained the on-treatment relapse hazard ratio from the largely overlapping meta-analysis by Zhou et al. (2020), which reports hazard ratios for relapse in maintenance/discontinuation randomized trials.

Specifically, we extracted the estimated hazard ratio at 6 months from Zhou et al. (2020)’s pooled survival-curve model (their Supplemental Appendix Figure 6) by digitizing the figure. We used the 6-month estimate because it is supported by many of the contributing trials, and because the point estimate was not meaningfully different at other reported months. We applied this as a constant multiplicative effect on relapse hazards in the model (hazard ratio at 6 months ≈ 0.47; 95% CI ≈ 0.42 to 0.53).

#### Because Zhou et al. (2020) pooled populations containing both chronic and non-chronic depressive courses, we treated the reported hazard ratio of 0.47 as a mixed-population effect rather than as the effect for non-chronic relapse alone. To recover the non-chronic component, we estimated that the chronic-course subgroup made up about 29% of the pooled population (derived from the chronic-course probability in Section I.C1 below) and assumed that maintenance treatment was half as effective at preventing relapse in chronic depression as in non-chronic depression (base-case assumption; Section I.C4). Under those assumptions, the implied hazard ratio for non-chronic relapse prevention is about 0.41, and the corresponding hazard ratio for chronic-course relapse prevention is about 0.64. In the simulation, the non-chronic hazard ratio is supplied directly, and the chronic-course hazard ratio is derived internally using the assumed attenuation. This preserves the observed mixed-population hazard ratio of 0.47 by construction. For calibration steps that target mixed-population benchmarks, we use the reported hazard ratio of 0.47 directly. In probabilistic sensitivity analysis, the mixed-population hazard ratio is decomposed separately within each draw using that draw’s sampled chronic-course probability, preserving the dependence between treatment effect and chronic-course parameters.

#### I.B. Acute Remission Parameters

##### I.B0) Overview

Remission timing can be informed by two complementary literatures that emphasize different parts of the episode course. Acute antidepressant trials typically report remission over short horizons (e.g., 6–8 weeks) and show substantial early improvement and drug–placebo separation. In contrast, epidemiologic cohort studies that follow patients after an index episode show a pronounced slowing of recovery over longer horizons, with a non-trivial fraction remaining depressed at one year and only modest additional decline thereafter. A monthly course model intended to support multi-year stop/restart decisions must reconcile these short-horizon trial patterns with longer-horizon persistence.

##### I.B1) Monthly remission probability off antidepressants (acute/non-chronic episodes).

***In brief:*** *The off-ADM acute remission probability was identified jointly with the chronic-course parameters by calibrating the model’s mixture to three targets simultaneously: Gibbons et al. (2012) 6-week placebo remission, and Spijker et al. (2002) 12- and 21-month persistence (p_remit,acute,off ≈ 0.314/month).*

**In detail:** We parameterized acute (non-chronic) remission off antidepressants using a constant monthly remission hazard. To anchor early remission under off-ADM conditions, we used the placebo-arm cumulative remission at 6 weeks of 29.3% from Gibbons et al. (2012). We used Gibbons et al. (2012) because it reports an absolute placebo remission proportion; more recent antidepressant meta-analyses emphasize relative effects or symptom-scale change and do not provide an equally direct absolute placebo remission target for this purpose. While placebo arms are not equivalent to “no treatment,” our off-ADM strategy is also not “no care”: it involves active monitoring and follow-up, plausibly capturing some of the non-specific supports present in trial placebo conditions.

For orientation, we first show the approximate monthly remission rates implied by the 6-week placebo target under a simplified version of the model. If one treats the placebo 6-week remission as arising from a constant remission hazard in a single homogeneous group, 0.293 remitting by 6 weeks corresponds to an average monthly remission probability of about 0.222. However, our model allows heterogeneity in episode course. Using the chronic-course quantities estimated in Section I.C (p_chronic ≈ 0.28; p_remit,chronic,off ≈ 0.014 per month), the same overall 6-week placebo remission implies an acute/non-chronic off-ADM monthly remission probability of approximately 0.31, because the faster-remitting acute episodes must offset the much slower-remitting persistent-course episodes in the mixture.

The way we implement this formally is to identify the acute/non-chronic off-ADM remission probability jointly with the chronic-course frequency and chronic off-ADM remission rate by calibrating the model’s off-ADM mixture outcomes to three targets: Gibbons et al. (2012) 6-week placebo remission (0.293), Spijker et al. (2002)’s 12-month unremitted proportion (0.24), and Spijker et al. (2002)’s 21-month unremitted proportion (0.20). Because these parameters are jointly calibrated to the same targets, they are not treated as independent in probabilistic analyses. We propagate uncertainty by sampling the three calibration targets within their reported 95% confidence intervals (Gibbons et al. (2012) 6-week placebo remission: 0.293; Spijker et al. (2002)’s 12-month unremitted proportion: 0.24, 95% CI 0.18–0.30; Spijker et al. (2002)’s 21-month unremitted proportion: 0.20, 95% CI 0.14–0.26) and re-solving the calibration system in each draw; the resulting empirical percentiles define the reported uncertainty intervals for the off-ADM acute/non-chronic remission probability.

##### I.B2) Remission hazard ratio on antidepressants (on vs off ADM).

***In brief:*** *The on-treatment remission hazard ratio was calibrated to reproduce Gibbons et al. (2012) 6-week drug-arm remission given the off-ADM parameters and ramp assumption (HR_remit ≈ 2.08).*

In detail: We modeled antidepressant effects on episode resolution as a multiplicative effect on the remission hazard. Because acute trials typically report remission as a fixed-time endpoint (rather than a time-to-event hazard ratio), we identified the on-treatment remission hazard ratio HR_remit by calibration: we solved for the constant hazard multiplier that makes the model reproduce the Gibbons et al. (2012) 6-week drug-arm remission target, given the off-ADM remission parameters from Section I.B1 and the month-1 ramp assumption (f_ramp = 0.64; Section I.B3). For parsimony, we applied the same hazard ratio to both acute/non-chronic and persistent-course episodes.

To represent uncertainty, we repeated this calibration using the lower and upper bounds of the Gibbons et al. (2012) 6-week drug-arm remission target (i.e., substituting the reported 95% CI bounds for the drug-arm remission proportion) and took the resulting calibrated HR values as the lower and upper limits for the hazard ratio.

##### I.B3) Treatment-effect ramp after initiation or restart (month 1).

***In brief:*** *We modeled month-1 treatment as partial-strength using Henssler et al. (2018)’s ratio of drug–placebo separation at 4 weeks versus 12 weeks, yielding a ramp fraction of ≈ 0.64 (full effect from month 2 onward).*

**In detail:** We modeled antidepressant benefit on remission as ramping up after initiation or restart rather than appearing at full strength immediately. Henssler et al. (2018) is a meta-analysis of acute antidepressant placebo-controlled trials focused on the time-course of antidepressant response. In Henssler et al. (2018), most of the increase in drug–placebo separation occurs by about month 2; the main deviation from full effect is in the first month. We therefore modeled month 1 as partial-strength treatment and treated months 2+ as full effect.

We parameterized the month-1 ramp fraction using Henssler et al. (2018), which reports pooled drug–placebo differences in continuous symptom severity over time. Let D(t) denote the pooled drug–placebo difference in symptom severity at week t, expressed as percent of baseline score. We defined the ramp fraction as f = D(4 weeks) / D(12 weeks), i.e., the fraction of the 12-week symptom-severity separation that is present by week 4. Using Henssler et al. (2018)’s estimates (D(4) = 7% and D(12) = 11%), this yields f ≈ 0.64, which we apply as the partial-strength treatment effect in month 1 (with full effect from month 2 onward).

To represent uncertainty in f, we propagated the reported 95% confidence intervals for D(4) (5%–9%) and D(12) (9%–13%) using log-scale error propagation for a ratio. We assumed a correlation of 0.8 between the week-4 and week-12 pooled estimates, consistent with common practice in robust variance estimation meta-analysis (e.g., the robumeta default) (37). This yields an approximate 95% uncertainty interval for the ramp fraction of 0.53–0.76.

#### I.C. Persistent Residual Symptoms Parameters

##### I.C0. Overview: chronic-course episodes and persistent residual symptoms

To reproduce the observed distribution of depressive episode durations in routine care, we represented heterogeneity in persistence using a two-course mixture. At the onset of each new episode, the episode is assigned to either a non-chronic course (typical resolution over months) or a chronic course (a slower, persistent course). Episodes assigned to the chronic course transition, after the acute phase, into a persistent residual-symptom state that can persist for extended periods and remit at a slower monthly rate (labeled “entrenchment” in Figure S1). This “chronic course” construct is a pragmatic modeling device to capture the long right tail of episode persistence and does not map one-to-one onto DSM chronic depression duration criteria. It is not intended to diagnose dysthymia or persistent depressive disorder; it represents persistent, lower-severity depressive morbidity after the acute phase of some episodes.

##### I.C1. Chronic-course mixture calibration (base chronic-course probability and chronic residual-state remission off ADM)

***In brief:*** *The chronic-course probability (≈ 29%) and chronic-state monthly remission rate (≈ 1.4%/month off ADM) were jointly calibrated with the acute remission parameters to reproduce both Gibbons et al. (2012) early remission and Spijker et al. (2002) long-tail persistence.*

In detail: This section parameterizes two quantities that govern chronic-course persistence: (1) the base probability that a new episode follows a chronic course (p_chronic) and (2) the monthly remission probability off antidepressant medication while in the persistent residual-symptom state (p_remit,chronic,off). These parameters are jointly identified by the long-duration tail of observed episode persistence and were therefore calibrated together rather than treated as independent inputs.

Because Spijker et al. (2002) reflects routine care rather than a standardized treatment protocol, the reported unremitted proportions at follow-up represent a mixture of time on and off antidepressant medication and a mixture of patient risk levels. To make the Spijker et al. (2002) persistence targets commensurate with our simulation outputs, we approximated (i) the fraction on antidepressants during the Spijker et al. (2002) persistence window and (ii) the risk-band distribution represented by the Spijker et al. (2002) sample, and then calibrated the chronic-course mixture so that simulated, mixture-averaged persistence matched Spijker et al. (2002).

For antidepressant exposure during the persistence window, we combined two quantities from the companion manuscript Spijker et al. (2001): 67.2% of participants received any professional care and 42.6% of care users received antidepressant medication (31). Their product (0.672 × 0.426 ≈ 0.29) was used as a single-point estimate of the proportion on antidepressants during follow-up, and persistence targets were evaluated under an on/off exposure mixture consistent with this estimate.

To approximate the Spijker et al. (2002) risk-band distribution, we mapped Spijker et al. (2002) clinical descriptors to our relapse-risk strata. Spijker et al. (2002) reports that 43.2% of episodes occurred among patients with recurrent depression and 30.4% were severe. Because the joint distribution of recurrence and severity was not reported, we assumed severity was twice as prevalent among recurrent than non-recurrent cases. Under this assumption, we allocated episodes across strata as follows: episodes that were neither recurrent nor severe were assigned to Very Low; severe but non-recurrent episodes to Low; recurrent non-severe episodes to Moderate; and recurrent severe episodes to High, yielding an implied distribution of 44.7% Very Low, 12.1% Low, 24.9% Moderate, and 18.3% High. In assigning these strata we relied on the fact that past recurrence is a greater predictor of future recurrence than is baseline severity.

We calibrated (p_chronic, p_remit,chronic,off) within the joint remission calibration described in Section I.B, using Spijker et al. (2002)’s unremitted proportions to anchor persistence beyond the acute treatment window while maintaining consistency with acute-phase placebo remission targets. In probabilistic sensitivity analysis, we preserved the induced correlation between these parameters using the joint-sampling approach from Section I.B: in each draw, we sampled the calibration targets within their reported uncertainty and re-solved the calibration, yielding paired draws of (p_chronic, p_remit,chronic,off) that jointly reproduce the persistence targets under the assumed antidepressant-exposure and risk-band mixtures. The resulting empirical percentiles define the uncertainty intervals reported in the parameter table.

##### I.C2. Burden weight of persistent residual symptoms

***In brief:*** *The burden weight for the persistent residual-symptom state (0.44 on a 0–1 scale) was derived from Judd et al. (1998)’s distribution of weekly symptom severity across four categories, mapped to proportional weights.*

**In detail:** We included a persistent residual-symptom state in the simulation and required a parameter for its burden per day. For outcome accounting, we expressed burden using a 0–1 weight, where 0 corresponds to full remission and 1 corresponds to a full major depressive episode as represented in the model.

We informed this weight using Judd et al. (1998), which reports long-term weekly symptom status across four mutually exclusive categories: healthy/asymptomatic, subsyndromal symptoms, minor depression, and full-syndromal major depression. This framework captures that patients with persistent depressive morbidity cycle over time rather than remaining continuously at full-syndromal severity.

To derive a single burden weight, we mapped Judd et al. (1998)’s four weekly categories to a 0–1 scale (healthy/asymptomatic = 0; subsyndromal symptoms = 1/3; minor depression = 2/3; full-syndromal major depression = 1) and computed the weighted mean of time spent across each category, which yielded a burden weight of 0.44.

Uncertainty was approximated by propagating sampling variability from the Judd et al. (1998) category means to the weighted mean; this produced an approximate 95% interval of 0.39 to 0.50 for probabilistic sensitivity analysis.

##### **I.C4. Chronic relapse prevention fraction**

***In brief:*** *Because direct evidence on differential relapse prevention by course type is sparse, we assumed maintenance antidepressants are half as effective at preventing chronic-course relapses as non-chronic relapses (base case 0.5; varied 0 to 1 in sensitivity analysis).*

In detail: We assumed that antidepressant maintenance reduces the risk of relapse, but that its relapse-preventive effect may be attenuated for the subset of relapses that would otherwise follow a chronic, persistent course. Conceptually, we treated relapses as a mixture of non-chronic and chronic-course components. Antidepressant maintenance was assumed to prevent non-chronic relapses with the full relapse-prevention effect used elsewhere in the model, but to have a weaker preventive effect on the chronic-course component. This reflects the clinical intuition that chronic-course episodes may arise in the setting of sustained stressors and entrenched vulnerabilities (38) that are less preventable by pharmacologic buffering alone.

Because we found little direct evidence to quantify differential prevention by relapse type, we introduced a chronic relapse prevention fraction to represent this structural uncertainty. Operationally, this fraction scales the antidepressant’s relapse-preventive effect for chronic-course relapses relative to non-chronic relapses. We set the base-case value to 0.5, indicating that maintenance antidepressants are assumed to be half as effective at preventing chronic-course relapses as they are at preventing non-chronic relapses. In the model, this is implemented as an attenuation of the preventive effect on the chronic component: the chronic-course relapse HR is exp(CREF × log(HR_nc)), where HR_nc is the decomposed non-chronic HR from Section A5 (e.g., with CREF = 0.5 and HR_nc ≈ 0.41, the chronic HR ≈ 0.64). We evaluated this assumption using scenario and one-way sensitivity analyses: 0 (no preventive effect for chronic-course relapses) and 1 (full preventive effect, equal to that for non-chronic relapses).

I.D. Model Structure Parameters

##### **I.D0. Overview: model structure parameters**

Section D specifies fixed structural inputs that define the simulated treatment strategies and follow-up window (continuation duration after remission, time horizon, and clinical monitoring frequency) and a reporting convention (temporal discounting). These quantities are primarily design choices chosen for clinical plausibility and interpretability, and we evaluate key alternatives in sensitivity analyses where appropriate.

I.D1. Continuation duration

We set the continuation phase to 8 months after remission. This duration is coherent with the model’s early post-remission period of elevated relapse risk and falls within commonly recommended continuation ranges (e.g., 6–12 months after remission). Because the continuation duration parallels this simplified hazard structure, we did not treat 6- or 12-month continuation as primary sensitivity analyses.

I.D2. Time horizon

We set a 5-year horizon to capture meaningful longer-term consequences of maintenance vs discontinuation without extrapolating far beyond what typical evidence can reasonably support. This also aligns with the availability of longer follow-up data in first-episode cohorts, including Bukh et al. (2016), which reports outcomes over 5 years. In sensitivity analyses where the time horizon was instead set to 3 years or 10 years, we tested the impact of this horizon choice.

I.D3. Clinical monitoring

Quarterly monitoring was chosen to operationalize discontinuation as an actively monitored strategy. We also considered monthly monitoring to explore the impact of close follow-up, and only a twice yearly follow-up.

I.D4. Temporal Discounting

We did not apply temporal discounting. Outcomes were expressed in patient-centered time units (days with depression and days of antidepressant exposure), without assigning monetary costs. Both the “costs” (medication exposure) and “benefits” (depressed days averted) accrue throughout the relatively bounded 5-year horizon, so discounting would be expected to affect both streams similarly (in contrast to settings where costs occur upfront and benefits occur much later). In addition, there is no clear consensus discount rate for these non-monetary outcomes. We therefore report undiscounted totals.

### II. Configuration of Verifications and Validations

##### Overview and principles

The goal of the verifications and validations is to assess whether the simulation reproduces the core clinical behaviors it is intended to represent—relapse/recurrence after remission (on vs off antidepressants), remission dynamics while depressed (including early treatment ramp-up), and longer-tail persistence.

These checks are not performed by rerunning the headline policy experiments unchanged. Instead, for each benchmark we run the same model machinery under a like-for-like setup that matches the structure of the target evidence as closely as possible—for example, using the appropriate mixture of risk strata, matching the relevant treatment contrast (maintenance vs discontinuation or on- vs off-treatment), and aligning the model’s “time since remission” or “time since episode onset” indexing to the way follow-up is defined in the source. This produces model outputs that correspond directly to the reported quantities in each benchmark.

We use “verification/validation” as a practical label for these benchmark comparisons rather than as a strict technical distinction. Because parameters were often set to reproduce broader empirical features rather than isolated datapoints, the boundary between directly constrained and out-of-fit comparisons is not always sharp. We therefore identify, for each benchmark, which features were used to set parameter values and which were left for comparison. Together, these checks assess whether the model reproduces the clinically relevant patterns it was designed to capture.

We compared the model against benchmarks for four core clinical features: (1) high-risk relapse over the first 12 months after remission, including on- versus off-maintenance differences; (2) lower-risk recurrence over multiple years; (3) the acute-phase remission level and the early ramp-in of antidepressant benefit after initiation; and (4) the long-tail persistence of depressive episodes over months to years (e.g., unremitted proportions).

##### II.A1. Relapse: high-risk 12-month trajectory

**Clinical feature and benchmark.** We compared the model’s high-risk relapse behavior against a 12-month benchmark for cumulative relapse at 3, 6, 9, and 12 months after randomization, in both the discontinuation/placebo (off-treatment) and maintenance (on-treatment) arms. The goal was to assess whether the model captures (i) the overall level of relapse risk in a high-risk population, (ii) the characteristic pattern of higher early relapse risk that settles to a later steady-state level, and (iii) the expected separation between maintenance and discontinuation trajectories. We did not benchmark against 15 and 18 months because the source authors noted these later estimates were less reliable due to sparse trial support.

**Calibration, degrees of freedom, and out-of-fit checks.** Within this benchmark, three tuning choices were specified from published summaries: (1) the high-risk late-period off-treatment relapse hazard level, identified from later follow-up in the Kishi et al. (2023) discontinuation/placebo trajectory; (2) a single early-post-remission multiplier (with a fixed transition pattern) chosen to reproduce the early-to-late shape of the Kishi et al. (2023) discontinuation/placebo trajectory; and (3) a single maintenance vs discontinuation hazard ratio for relapse prevention, taken from Zhou et al. (2020)’s pooled estimate at 6 months and applied as a constant multiplicative effect. We did not tune the model to match the individual cumulative relapse points at 3, 6, 9, and 12 months in either arm; instead, after setting these tuning choices, we compared the model-implied discontinuation and maintenance trajectories to the full set of reported timepoints as trajectory checks, recognizing that the timepoints within each arm are correlated and that the trial evidence base overlaps across summaries.

**Like-for-like simulation setup.** We simulated a cohort starting in remission, because the benchmark reports relapse after remission. We used the high-risk stratum to match the clinical severity mix represented by this benchmark and to align with how the high-risk relapse module was anchored. We generated two trajectories: an off-treatment trajectory corresponding to discontinuation/placebo, and an on-treatment trajectory corresponding to maintenance, implemented by applying a constant maintenance vs discontinuation relapse hazard ratio to the underlying off-treatment hazards. To align the simulation’s remission-based clock to the benchmark’s follow-up window, we assumed participants had already completed approximately two months of continuation treatment; accordingly, we began counting relapses and reading out outcomes starting two months after remission, and extracted cumulative relapse at 3, 6, 9, and 12 months.

##### II.A2. Relapse/recurrence: lower-risk multi-year trajectory

Clinical feature and benchmark. We compared the model’s lower-risk long-horizon recurrence behavior against a multi-year benchmark of cumulative recurrence following a first lifetime depressive episode (26). The goal was to assess whether the model reproduces (i) the overall low recurrence level expected after a first episode, (ii) separation between a very-low versus low recurrence stratum consistent with the benchmark’s severity split (mild vs moderate-to-severe), and (iii) the shape of cumulative recurrence over years 1–5 under these lower-risk dynamics.

**Calibration, degrees of freedom, and out-of-fit checks.** Within this benchmark, two tuning choices were fixed. First, we solved for the Very low-risk late-period off-treatment hazard so that the model’s pooled two-year cumulative recurrence—computed as the benchmark-weighted mixture of Very low (mild) and Low (moderate-to-severe) strata—matches the benchmark’s reported two-year recurrence (after accounting for assumed post-remission medication exposure). Second, we fixed the Low-risk hazard as a proportional scaling of the Very low-risk hazard using the benchmark’s reported recurrence hazard ratio for moderate-to-severe versus mild. After these were set, the remaining benchmark horizons (years 1 and 3–5) served as out-of-fit checks of the multi-year trajectory.

**Like-for-like simulation setup.** We simulated cohorts starting in remission and followed them for five years, with two strata corresponding to the benchmark’s severity split (mild mapped to Very low-risk; moderate-to-severe mapped to Low-risk) and pooled results computed using the benchmark’s observed proportions (≈24% mild, ≈76% moderate-to-severe). Because the benchmark does not standardize antidepressant exposure after remission, we imposed an explicit accounting assumption to translate the observed recurrence into off-treatment hazards: we assumed guideline-concordant continuation treatment for eight months after remission followed by discontinuation. We aligned the start of recurrence accumulation to the benchmark’s remission requirement by beginning outcome counting after the initial two-month remission period (i.e., treating the first modeled follow-up month as post-remission month 3 when reading out years 1–5 cumulative recurrence).

##### II.B. Acute remission level and early time-course

**Clinical feature and benchmark.** We compared the model’s acute-phase remission behavior against two complementary benchmarks. First, we used Gibbons et al. (2012) to assess the absolute level of early remission on and off antidepressants at a short fixed horizon (6 weeks). Second, we used Henssler et al. (2018) to assess the early time course of antidepressant benefit using normalized, within-study quantities. Specifically, when comparing the model with Henssler et al. (2018), we treated the 12-week on-antidepressant remission level as 100%, and compared (in both the benchmark and the model) the corresponding percentages for 4-week and 8-week remission on antidepressants, 4-week and 8-week remission off antidepressants, and 12-week remission off antidepressants.

**Calibration, degrees of freedom, and out-of-fit checks.** The absolute acute-remission behavior was anchored using the two Gibbons et al. (2012) 6-week remission targets (placebo and drug). In the current model, these acute targets are satisfied in the presence of latent heterogeneity in course, because early remission outcomes reflect a mixture of (i) acute/non-chronic episodes and (ii) episodes that transition into a persistent residual-symptom pathway; accordingly, the Gibbons et al. (2012) anchoring is performed jointly with the chronic-course frequency and the chronic residual-symptom remission rate (parameterized in the persistence section) so that the model reproduces early remission while also matching longer-horizon persistence. The time-course component uses Henssler et al. (2018)’s normalized pattern as the primary benchmark: we compare the model’s normalized remission percentages (each expressed relative to the model’s own 12-week on-antidepressant remission) to the corresponding Henssler et al. (2018) normalized percentages. Rather than tuning the model to match each normalized timepoint separately, the ramp specification imposes a simple structure (partial-strength effect in month 1; full strength from month 2 onward) and therefore allows the remaining normalized timepoints to function as trajectory checks of whether this structure reproduces Henssler et al. (2018)-like timing.

**Like-for-like simulation setup.** For both benchmarks, we simulated incident episodes using the same episode-course machinery as in the main analyses (including the latent mixture of non-chronic episodes and persistent-course episodes). To evaluate the Gibbons et al. (2012) targets, we ran two conditions—off antidepressants and on antidepressants—and read out cumulative remission at 6 weeks. To evaluate the Henssler et al. (2018) time-course benchmark, we ran the same two conditions and read out cumulative remission at 4, 8, and 12 weeks, then normalized all quantities by setting the model’s 12-week on-antidepressant remission to 100% and expressing the remaining five quantities (4/8 weeks on; 4/8/12 weeks off) as percentages of that reference. We used a 1-week model time-step for this verification/validation because this allowed us to more precisely match the 6-week endpoint.

##### II.C. Episode persistence

**Clinical feature and benchmark.** We compared the model’s long-tail episode persistence against Spijker et al. (2002), which reports the proportion of participants unremitted at 3, 6, 12, and 21 months after an index episode. The goal was to assess whether the model reproduces the clinically important slowing of recovery over longer horizons, including a non-trivial fraction remaining depressed at one year with only modest additional decline thereafter.

**Calibration, degrees of freedom, and out-of-fit checks.** Persistence beyond the acute window is generated in the model by a two-course mixture: some episodes follow an acute/non-chronic course, while others transition into a persistent residual-symptom pathway with slower remission. Two quantities governing this persistent pathway—the overall chronic-course probability and the monthly remission probability in the persistent residual-symptom state off antidepressants—were calibrated using the Spijker et al. (2002) persistence targets, jointly with the acute-placebo remission anchoring described in the acute-remission section. The 12- and 21-month unremitted proportions served as the primary persistence anchoring features in this joint calibration, while the earlier 3- and 6-month proportions were compared as out-of-fit checks of whether the resulting persistence curve also matches intermediate horizons without additional tuning.

**Like-for-like simulation setup.** For this benchmark, we simulated cohorts starting in an active depressive episode at episode onset and followed them forward to read out the proportion unremitted at 3, 6, 12, and 21 months. To make the Spijker et al. (2002) targets commensurate with simulation outputs, we evaluated persistence under a fixed mixture of time on versus off antidepressants during follow-up rather than a single exposure state, using the same accounting assumption as in parameter derivation (a single-point estimate of the proportion on antidepressants during the persistence window). We also computed persistence as a pre-specified mixture across relapse-risk strata using the Spijker et al. (2002)-mapped risk-band distribution described in parameter derivations, so that the simulated non-remittance curve reflects the benchmark’s implied mix of severity and recurrence risk.

##### II.D. Verification and Validation Key Numerical Comparisons

Table S2 presents the key numerical comparisons underlying the verification and validation panels shown in Figure 2 of the main text. For each benchmark, it reports the published estimate (with 95% confidence interval where available), the corresponding model estimate, and the absolute difference in percentage points, along with the calibration status of each comparison as described above.

**Table S2. Key numerical comparisons for model verification and validation**

| **Arm** | **Timepoint** | **Published (95% CI)** | **Model** | **Diff (pp)** |
| --- | --- | --- | --- | --- |
| **Cumulative relapse: High-risk 12-month trajectory (Kishi et al. 2023) — Panel A** | | | | |
| Placebo | 3 mo | 22.1% (17.9, 25.7) | 24.5% | +2.4 |
| Placebo | 6 mo | 35.6% (31.0, 39.6) | 34.3% | −1.3 |
| Placebo | 9 mo | 39.8% (34.3, 45.3) | 38.7% | −1.1 |
| Placebo | 12 mo | 44.1% (35.9, 51.6) | 42.9% | −1.2 |
| Maintenance | 3 mo | 10.8% (8.8, 13.2) | 13.0% | +2.2 |
| Maintenance | 6 mo | 17.3% (14.6, 20.3) | 18.5% | +1.2 |
| Maintenance | 9 mo | 18.7% (14.8, 23.4) | 21.4% | +2.7 |
| Maintenance | 12 mo | 21.6% (15.9, 28.8) | 23.9% | +2.3 |
| **Cumulative recurrence: Lower-risk multi-year trajectory (Bukh et al. 2016) — Panel B** | | | | |
| Weighted | 1 yr | 9.0% (5.4, 12.6) | 8.7% | −0.3 |
| Weighted | 2 yr | 15.2% (10.7, 19.7) | 15.3% | +0.1 |
| Weighted | 3 yr | 20.5% (15.5, 25.5) | 21.2% | +0.7 |
| Weighted | 4 yr | 24.8% (19.4, 30.2) | 26.8% | +2.0 |
| Weighted | 5 yr | 31.5% (25.7, 37.3) | 32.0% | +0.5 |
| **Cumulative remission: Absolute level (Gibbons et al. 2012) — Panel C** | | | | |
| Placebo | 6 wk | 29.3% (27.8, 30.8) | 29.5% | +0.2 |
| Maintenance | 6 wk | 43.0% (41.6, 44.4) | 42.1% | −0.9 |
| **Cumulative remission: Normalized time-course (Henssler et al. 2018) — Panel C*** | | | | |
| Maintenance | 4 wk | 46.5% (30.2, 60.5) | 49.0% | +2.5 |
| Maintenance | 8 wk | 79.1% (67.4, 88.4) | 82.1% | +3.0 |
| Placebo | 4 wk | 20.9% (11.6, 30.2) | 33.4% | +12.5 |
| Placebo | 8 wk | 53.5% (27.9, 81.4) | 57.7% | +4.2 |
| Placebo | 12 wk | 69.8% (37.2, 100.0) | 74.8% | +5.0 |
| **Episode persistence: Long-tail trajectory (Spijker et al. 2002) — Panel D** | | | | |
| Mixed | 3 mo | 50% (44, 56) | 46.0% | −4.0 |
| Mixed | 6 mo | 37% (31, 43) | 31.4% | −5.6 |
| Mixed | 12 mo | 24% (18, 30) | 23.9% | −0.1 |
| Mixed | 21 mo | 20% (14, 26) | 20.0% | 0.0 |

* Henssler et al. (2018) values are expressed as % of 12-week on-antidepressant remission (= 100% reference); 12-week maintenance row omitted as it is the normalization anchor by definition.

### III. Results of Univariate Sensitivity Analyses

Values are medication-years per depression-month averted (point estimates). Probabilistic uncertainty is presented in Table 2 and Figure 3 of the main text. Base-case values may differ slightly from Table 2 because sensitivity analyses use fixed parameters rather than the median across probabilistic draws.

Table S3. Univariate sensitivity analyses

| **Monitoring frequency sensitivity** | | | |
| --- | --- | --- | --- |
| **Risk band** | **Monthly** | **Quarterly (base)** | **Every 6 months** |
| Very low | 13.3 | 11.4 | 10.2 |
| Low | 5.9 | 5.1 | 4.5 |
| Moderate | 3.2 | 2.7 | 2.4 |
| High | 1.7 | 1.4 | 1.3 |
| **Time-horizon sensitivity** | | | |
| **Risk band** | **36 months** | **60 months (base)** | **120 months** |
| Very low | 13.4 | 11.5 | 9.8 |
| Low | 5.9 | 5.1 | 4.3 |
| Moderate | 3.2 | 2.7 | 2.3 |
| High | 1.7 | 1.4 | 1.2 |
| **Chronic-prevention sensitivity** | | | |
| **Risk band** | **None** | **Half (base)** | **Full** |
| Very low | 20.4 | 11.5 | 9.0 |
| Low | 8.7 | 5.1 | 4.0 |
| Moderate | 4.5 | 2.7 | 2.2 |
| High | 2.3 | 1.4 | 1.2 |

### Supplemental References

35. Kessler RC, Berglund P, Demler O, Jin R, Merikangas KR, Walters EE. Lifetime prevalence and age-of-onset distributions of DSM-IV disorders in the National Comorbidity Survey Replication. Arch Gen Psychiatry. 2005;62(6):593-602. doi:10.1001/archpsyc.62.6.593

36. Kessler RC, Chiu WT, Demler O, Merikangas KR, Walters EE. Prevalence, severity, and comorbidity of 12-month DSM-IV disorders in the National Comorbidity Survey Replication. Arch Gen Psychiatry. 2005;62(6):617-627. doi:10.1001/archpsyc.62.6.617

37. Fisher Z, Tipton E. robumeta: An R-package for robust variance estimation in meta-analysis [Internet]. arXiv; 2015 [cited 2026 Mar 30]. Available from: http://arxiv.org/abs/1503.02220 doi:10.48550/arXiv.1503.02220

38. Dougherty LR, Klein DN, Davila J. A growth curve analysis of the course of dysthymic disorder: the effects of chronic stress and moderation by adverse parent-child relationships and family history. J Consult Clin Psychol. 2004 Dec;72(6):1012-21. doi:10.1037/0022-006X.72.6.1012 PubMed PMID: 15612848.
